# Early Syndecan-4 Upregulation Predicts Cognitive and Pathological Trajectories in Alzheimer Disease

**DOI:** 10.1101/2025.07.22.25331841

**Authors:** Rawan Tarawneh, Krista L. Moulder, Danai G. Topouza, Amrita Kar, Heng (Tony) Qian, Vernon S. Pankratz, Jigyasha Timsina, Erin E. Franklin, Suzanne E. Schindler, Richard J. Perrin, Beau M. Ances, Tammie L.S. Benzinger, Jason Hassenstab, John C. Morris, David M. Holtzman, Carlos Cruchaga

**Affiliations:** Department of Neurology, Saint Louis University, St Louis, Missouri, USA 63110; Department of Neurology, Washington University in St Louis, St Louis, Missouri, USA 63110; Knight Washington University Alzheimer Disease Research Center, Washington University in St Louis, St Louis, Missouri, USA 63110; Olink Proteomics, Part of Thermo-Fisher Scientific, Boston, Massachusetts, USA 02453; Department of Internal Medicine, Division of Epidemiology, Biostatistics, and Preventive Medicine, University of New Mexico, Albuquerque, New Mexico, USA 87110; Department of Psychiatry, Washington University in St Louis, St Louis, Missouri, USA 63110; Neuro-Genomics and Informatics Center, Washington University in St Louis, St Louis, Missouri, USA 63110; Department of Pathology and Immunology, Washington University in St Louis, St Louis, Missouri, USA 63110; Hope Center for Neurological Disorders, Washington University in St Louis, St Louis, Missouri, USA 63110; Department of Radiology, Washington University in St Louis, St Louis, Missouri, USA 63110; Department of Psychological and Brain Sciences, Washington University in St Louis, St Louis, Missouri, USA 63110

**Keywords:** syndecan-4 (SDC4), Alzheimer disease, brain endothelial dysfunction, amyloid, tau, preclinical Alzheimer disease, cognition, disease trajectories, longitudinal cohort, pseudo-time

## Abstract

**Objective:** Brain endothelial dysfunction is an early pathological feature of Alzheimer disease (AD). Syndecan-4 (SDC4) is a heparan sulfate proteoglycan which is an important component of the brain endothelial glycocalyx. In this study, we investigate SDC4 associations with amyloid and tau pathologies and cognitive impairment in a large longitudinal cohort of AD, including preclinical AD, and controls.

**Methods:** The study included *n*=1,041 (*n*=802 cognitively unimpaired and *n*=239 cognitively impaired) participants. Biological classification using the National Institute of Aging-Alzheimer’s Association “ATN” framework was performed in all participants. Cognitive assessments included the Clinical Dementia Rating^®^-sum of boxes and the Knight-Preclinical Alzheimer’s Cognitive Composite which includes episodic memory, language, and executive function scores. Cerebrospinal fluid (CSF) total tau, p-tau181, and Aβ42/Aβ40 levels were measured using Lumipulse assays. CSF measures of SDC4 and established AD biomarkers were obtained using the Explore HT Olink Proteomics platform. Amyloid-PET (*n*=719) and tau-PET (*n*=302) scans were performed in subsets of participants. Partial correlations and linear mixed models, respectively, examined cross-sectional and longitudinal associations of CSF SDC4 levels with amyloid-PET and tau-PET burden and cognition adjusting for co-variates. CSF biomarker trajectories across the course of AD progression were estimated using pseudo-time models.

**Results:** CSF SDC4 levels were elevated in AD, including the earliest preclinical stages, compared to controls and were closely associated with CSF and imaging biomarkers of amyloid and tau, and CSF biomarkers of neuronal and synaptic injury, astrocytic reactivity and microglial dysregulation. Higher CSF SDC4 levels correlated with higher global and regional amyloid-PET and tau-PET burden and worse baseline cognition. Higher baseline CSF SDC4 levels predicted more rapid progression of brain amyloid and tau, and faster decline in global cognition, episodic memory, language, and executive functions over a mean follow-up period of 8 years. SDC4 associations with cognition were mainly mediated by global tau-PET burden. Importantly, our pseudo-time models estimate that SDC4 upregulation begins very early in AD pathogenesis near the point of amyloid-positivity and increases more robustly following the point of tau-positivity. In our cohort, SDC4 was among the top 10 most important proteins (of >5,000 examined proteins) in predicting the pseudo-time models of AD progression and predicted these models to a potentially better extent than established AD biomarkers.

**Interpretation:** Findings from this large translational longitudinal study suggest close associations of SDC4 upregulation with the progression of brain amyloid and tau pathologies in AD including the early preclinical stages. Further, we here demonstrate the ability of early increases in CSF SDC4 levels to predict trajectories of future cognitive decline over a mean follow-up period of 8 years. Therefore, we propose that SDC4 upregulation, as a potential surrogate of brain endothelial dysfunction, is an early event in AD pathogenesis which can reliably predict cognitive and pathological disease trajectories.

## 1. Introduction

Brain endothelial dysfunction refers to characteristic structural or functional alterations that occur at the intracellular or cell membrane level and interfere with endothelial cell integrity or function.^1–6^ Brain endothelial dysfunction is a common pathology which is observed in almost all (>90%) Alzheimer disease (AD) brains, including those with only mild AD pathology, and occurs independently of alterations to other vascular constituents and the presence or severity of other forms of vascular pathology (e.g., atherosclerosis or arteriolosclerosis).^5,6^

Emerging evidence from clinical and animal studies supports an important role for brain endothelial dysfunction in early AD pathogenesis, including the onset or progression of pathologic amyloid and tau.^7–10^ Human transcriptomics studies suggest that endothelial pathways are among the most differentially expressed in AD brains;^11^ 30 of the top 45 genes associated with AD were found to be expressed in vascular cell types, and several of these had their highest expression levels in the brain endothelium.^12^ Endothelial dysfunction displays regional and laminar patterns that parallel those of neuronal loss^3^ and is preferentially localized in the vicinity of dystrophic neurites.^3,13^ Further, endothelial dysfunction precedes amyloid deposition^8^ and cognitive deficits in AD transgenic and insulin-resistant mice.^14^ Despite its prevalence, the role of brain endothelial dysfunction in AD pathogenesis and its relation to cognitive impairment and AD pathology have not been adequately examined in clinical cohorts due to the absence of reliable endothelial biomarkers.^15^

Syndecan-4 (SDC4) is an abundant cell-surface heparan sulfate proteoglycan (HSPG) which is part of the brain endothelial glycocalyx and plays an important role in regulating endothelial cell migration, proliferation, and signaling, and mediating endothelial interactions with the extracellular matrix and various growth factors (e.g., vascular-endothelial growth factor).^16–19^ Upregulation of SDC4 expression has consistently been demonstrated across different regions in human AD brains.^20,21^ However, the relationship between SDC4, AD pathology, and cognition has not been adequately investigated in cross-sectional or longitudinal AD cohorts. In this study, we aim to investigate the role of brain endothelial dysfunction in AD by characterizing the relationship between cerebrospinal fluid (CSF) SDC4 levels, as a potential surrogate of brain endothelial dysfunction, and the onset or progression of amyloid and tau pathologies, cognitive decline, and established AD biomarkers.

We here employ advanced proteomics in a large well-characterized longitudinal cohort of AD and controls (*n*=1,041), who have been followed for an average of 8 years, and for whom we have detailed demographic, genotypic, and CSF biomarker measures and longitudinal clinical, cognitive, and PET imaging assessments, to conduct the first comprehensive in-depth translational longitudinal study of CSF SDC4 as a potential marker of brain endothelial dysfunction in AD. Specifically, we investigate associations of CSF SDC4 with the severity of brain Aβ plaque and neurofibrillary tangle pathologies and their longitudinal progression over time. Further, we characterize the relationship between CSF SDC4, AD pathology, and cognition in participants across the AD continuum, including preclinical AD. Importantly, we use pseudo-time models to infer the trajectories of CSF SDC4 and established AD biomarkers over the course of AD progression and their temporal proximity to the onset of amyloid and tau pathologies.

Together, findings from this study highlight an important role for SDC4 upregulation in early AD pathogenesis, including its close association with the onset and progression of amyloid and tau pathologies across the AD continuum and its ability to reliably predict trajectories of future cognitive decline, even in the earliest preclinical stages. Importantly, our models suggest that SDC4 upregulation, as a surrogate of brain endothelial dysfunction, begins in close temporal proximity, and possibly prior, to significant amyloid deposition and predicts the pseudo-time models, which depict progression of participants along the clinicopathological stages of AD, to a potentially better extent than established AD biomarkers.

## 2. Methods

### 2.1. Study participants

Participants (*n*=1,041) were community-dwelling volunteers enrolled in longitudinal studies of healthy aging and dementia at the Knight Washington University Alzheimer Disease Research Center (Knight ADRC). Participants were in good general health with no other medical illness that could contribute to dementia and no contraindication to lumbar puncture (LP) or magnetic resonance imaging (MRI). The Clinical Dementia Rating (CDR^®^) was used to assess the presence and severity of cognitive impairment.^22^ A CDR 0 designation at baseline indicates normal cognition while a CDR ≥ 0.5 designation indicates cognitive impairment. Baseline clinical and cognitive assessments were those closest to the time of CSF collection (mean 3.2 months, median 2.7 months). Apolipoprotein E (*APOE*) genotypes were obtained in all participants as described.^23^ Participants with a history of traumatic brain injury, symptomatic cerebrovascular disease (i.e., stroke or intracranial hemorrhage), or significant small vessel disease on imaging (Fazekas ≥ 2) were excluded.

### 2.2. Cognitive assessments

All participants (*n*=1,041) had baseline CDR and CDR-sum of boxes (CDR-SB) assessments. Almost all participants (*n*=1,018) had baseline, and a large subset (*n*=898) had at least one follow-up, detailed 1.5-hour cognitive assessment which included measures of episodic memory, language, executive, and visuospatial functions and processing speed.^24^ Participants were followed for a mean ± standard error (SE) of 8.0 ± 0.16 years (median, 7.0 years).

The Preclinical Alzheimer’s Cognitive Composite (PACC) is a continuous measure of global cognition, which is sensitive to change in preclinical AD, and has been implemented in various preclinical AD cohorts with slight variations in calculation methods across different centers.^25,26^ The Knight-PACC used in this study^26^ was measured as a composite of the standardized sub-scores of four tests: episodic memory (the free recall score from the Free and Cued Selective Reminding Test [FCSRT-Free]),^27^ executive functions (Digit-Symbol Substitution test of the Weschler Adult Intelligence Scale-Revised^28^ and Trail-Making Part B [Trails-B]),^29^ and language (category fluency for animals [Animal Fluency]).^30^ The Knight-PACC was developed to increase sensitivity in preclinical AD and improve the psychometric characteristics of a composite by excluding measures with large ceiling or practice effects.^26^

### 2.3. CSF collection, storage, and processing

CSF samples (20–30 ml) were collected from all participants and analyzed for total tau, tau phosphorylated at threonine 181 (p-tau181), Aβ42, and Aβ40 levels (Lumipulse G1200, Fujirebio).^31^ CSF biomarker levels of SDC4, visinin-like protein-1 (VILIP-1), neurofilament-light chain (NFL), neurogranin (Ng), synaptosomal-associated protein-25 (SNAP-25), synaptotagmin-1 (SYT-1), glial fibrillary acidic protein (GFAP), chitinase 3-like protein-1 (YKL-40), and soluble triggering receptor expressed on myeloid cells-2 (sTREM2) were measured in all participants using the Explore HT proteomics panel (Olink Proteomics, part of ThermoFisher Scientific). Olink Proteomics is a highly sensitive and specific validated platform which utilizes antibody-based Proximity Extension Assay technology,^32^ and has demonstrated higher sensitivity, specificity and reproducibility compared to traditional antibody-based or mass-spectrometry approaches.^33^

### 2.4. Biological classification of study participants

All participants in this study (*n=*1,041) were classified according to the National Institute of Aging-Alzheimer’s Association (NIA-AA) “ATN” framework^34^ using the following cutoff values: Aβ+ defined as CSF Aβ42/Aβ40 <0.0673 and T+ defined as CSF p-tau181 >51.8 pg/ml (*n*=1,032). These cutoff values were chosen based on their ability to provide the highest predictive value for amyloid- and tau-PET positivity, respectively, in similar Knight-ADRC cohorts.^31^ Amyloid-PET and tau-PET data was used for AT classification in *n*=9 participants for whom CSF Aβ42/Aβ40 and p-tau181 levels were not available. Cognitively unimpaired individuals who had no biomarker evidence of amyloid or tau pathologies (i.e., CDR 0 A-T-) were defined as biomarker-confirmed healthy controls (*n*=539).

### 2.5. In vivo amyloid imaging

In vivo brain amyloid-PET imaging using the amyloid ligand Pittsburgh Compound-B (PiB; *n*=528) or 18F-AV-45 (Avid Radiopharmaceuticals, *n*=191) was obtained in a subset of participants (*n*=719) using the standard protocol (30–60 min postinjection as the time window for PiB and 50–70 min for 18F-AV-45).^35–37^ FreeSurfer calculations of partial volume-corrected SUVRs were obtained using the cerebellum as a reference region.^37^ The global amyloid burden was estimated using two measures: *i)* the mean cortical standardized uptake value ratio (SUVR), which is the average of the partial volume-corrected SUVRs from the bilateral precuneus, prefrontal cortex (superior frontal and rostral middle frontal), gyrus rectus (medial and lateral orbitofrontal) and lateral temporal (superior and middle temporal) regions^36^ and *ii)* Centiloid Units.^38^ The Knight ADRC has developed and validated equations for harmonization of amyloid PET data between amyloid tracers based on Deming regression and linear transformation in a calibration cohort.^39^ The following equation is used to harmonize partial volume-corrected mean cortical SUVR measures between PiB and 18F-AV-45: PiB SUVR= (18F-AV-45 SUVR−0.0238)/0.7948. PET-amyloid positivity is defined using a cutoff value for mean cortical (PiB) SUVR of ≥1.42.^39^ SUVR values from this global summary (without partial volume correction) were also converted into Centiloid Units to harmonize tracer and data processing differences using previously published equations.^38^

### 2.6. In vivo tau imaging

In vivo brain tau-PET imaging was performed using 18F-AV-1451 (Avid Radiopharmaceuticals) in a subset of participants (*n*=302) using the standard protocol.^40^ The standard procedure for tau-PET processing is identical to amyloid-PET processing except for the time window used for quantification (80-100 minutes for 18F-AV-1451 tau-PET). The global tau burden was estimated using a summary measure which was calculated by averaging the partial volume-corrected SUVRs from the bilateral amygdala, entorhinal, inferior temporal, and lateral occipital cortices.^41^ The proposed tau-PET positivity cutoff in the Knight ADRC is 1.22, using the above estimate of global tau burden and the cerebellum as a reference region. Braak staging is used to determine the progression of AD-related tau tangle pathology as described.^42^ The six stages of tau-PET progression are estimated using the following regional SUVRs: Braak 1-2 (entorhinal), Braak 3-4 (amygdala, nucleus accumbens, hippocampus, insula, medial and lateral orbitofrontal, pars orbitalis, parahippocampal, rostral and caudal anterior cingulate, posterior cingulate, and isthmus of the cingulate), and Braak 5-6 (banks of the superior temporal sulcus, middle temporal, caudal middle frontal, fusiform, inferior parietal, inferior temporal, lateral occipital, supramarginal and precuneus).

### 2.7. Statistical analysis

Student’s *t* tests, analysis of variance (ANOVA) and covariance (ANCOVA), Kruskal-Wallis, Fisher’s exact, or χ^2^ tests assessed differences in demographic, clinical, cognitive, genotypic, CSF biomarker or imaging measures between the study groups. Analyses were adjusted for multiple comparisons using false-discovery rate (FDR) corrections. Pearson correlations examined associations of CSF SDC4 with other CSF biomarkers. Partial correlations evaluated the relationship between CSF SDC4 levels and global or regional amyloid-PET (adjusting for age, sex, and *APOE* ε4 carrier status [*APOE* ε*4*]) and tau-PET (adjusting for age and sex) burden and cognition (i.e., CDR-SB, Knight-PACC, FCSRT-Free, Animal Fluency, Digit-Symbol, and Trails-B; adjusting for age, sex, education, and *APOE* ε*4*) (IBM SPSS Statistics v29.0.2). Mediation analyses examined whether global amyloid- or tau-PET burden mediated baseline associations of CSF SDC4 with cognition (adjusting for age, sex, education, and *APOE* ε*4*) using bootstrapping methods (SPSSv29.02, Process v4.1 by Hayes).^43^

Linear mixed models (PROC MIXED, SAS version 9.4; SAS Institute Inc), that specified a random subject-specific intercept and a random subject-specific slope, assessed the ability of baseline CSF SDC4 levels, examined as a continuous or categorical (i.e., divided into quartiles or terciles) variable, to predict annual change in cognition (adjusting for age, sex, education, and *APOE* ε*4*) and annual change in global amyloid-PET (adjusting for age, sex, and *APOE* ε*4*) or tau-PET (adjusting for age and sex) burden over follow-up. In these models, the β estimates (and *p* values) for the CSF SDC4*time interaction were used to determine whether baseline CSF SDC4 levels predicted annual rates (i.e., slopes) of progression in global amyloid- or tau-PET burden or cognitive decline over time. The time interval between baseline amyloid-PET (median interval, 2.1 months) or tau-PET (median interval, 6.5 months) scans and CSF collection were not significant predictors of baseline or longitudinal associations of CSF SDC4 with imaging outcomes and therefore, were not included in the final models.

Demographic, cognitive, CSF, and PET-imaging measures were standardized to *z* scores prior to analysis. Baseline cognitive, amyloid-PET and tau-PET assessments were those closest to the time of CSF collection. Statistical significance was defined as *p*<0.05.

#### 2.7.1. Pseudo-time Models

To generate the pseudo-time models, we inferred trajectories of CSF SDC4 and established AD biomarkers over a granular estimate of AD progression by creating a pseudo-time variable based on CSF biomarker levels of the core AD pathologies (CSF Aβ42/Aβ40 and CSF p-tau181) as described.^44^ Principal component analyses (PCA) generated two PCs which were included as input to a spectral embedding analysis using the igraph v2.1.1 package in R.^45,46^ Spectral embedding reduced the data nonlinearly to a latent two-dimensional space into which participants were embedded based on their biomarker values. We used the SCORPIUS v1.0.8 package in R to infer CSF biomarker trajectories in an unsupervised manner, which partitioned individual datapoints into clusters and optimized the shortest and smoothest path going through the center of these clusters.^47–49^ Therefore, each participant is assigned a coordinate along the inferred trajectory, in which 0 represents the lowest, and 1 the highest, level of pathology. Our pseudo-time model depicts the progression of participants sequentially through the following disease stages: CDR 0 A-T-(i.e., biomarker-confirmed healthy controls), CDR 0 A+T-(i.e., cognitively unimpaired participants with only amyloid pathology), CDR 0 A+T+ (i.e., biomarker-confirmed preclinical AD), and CDR ≥ 0.5 A+T+ (i.e., biomarker-confirmed symptomatic AD). The CSF biomarkers of interest were then plotted against the pseudo-time using smoothed generalized additive model lines from the ggplot2 v3.5.1 R package to estimate trajectories of biomarker changes across the disease course.^50^ CSF biomarker levels (as normalized protein expression [NPX] values reported on a log2 scale) were adjusted for mean protein levels (by centering to the median), and for age and sex using the “empiricalBayesLM()” function from the WGCNA v1.72-5 package in R.^51^ Adjusted biomarker log values were standardized to *z* scores prior to analysis.

We then used a random forest model implemented in the “gene_importances()” function of the SCORPIUS R package to rank the importance of each protein in predicting the ordering of participants along the pseudo-time model. In this model, feature importance is the mean decrease in mean squared error each time a feature is used in a decision tree split. This analysis used 10,000 trees over 10 permutations (1000 trees per permutation), with each tree sampling 1% of features. We calculated empirical *p*-values using permutation testing (10,000 permutations), with the addition of a pseudo-count of 1 to avoid *p* values of zero.^52^ *P* values were adjusted for multiple comparisons using the Benjamini & Hochberg procedure.

#### 2.7.2. Functional pathway analyses

Functional pathway analyses for SDC4 in human brains were performed using STRING v11.5 for functional protein association networks (https://string-db.org).^53^ (see **Supplement**)

## 3. Results

### 3.1. Demographics and cohort characteristics

A summary of the demographic, genotypic, clinical, cognitive, CSF biomarker, amyloid- and tau-PET imaging measures of the main study cohort and sub-cohorts are summarized in **Table 1 and Supplementary Table 1**. Most study participants (*n*=988) were CDR 0-0.5 (i.e., cognitively unimpaired or with mild cognitive impairment).

**Table 1.**
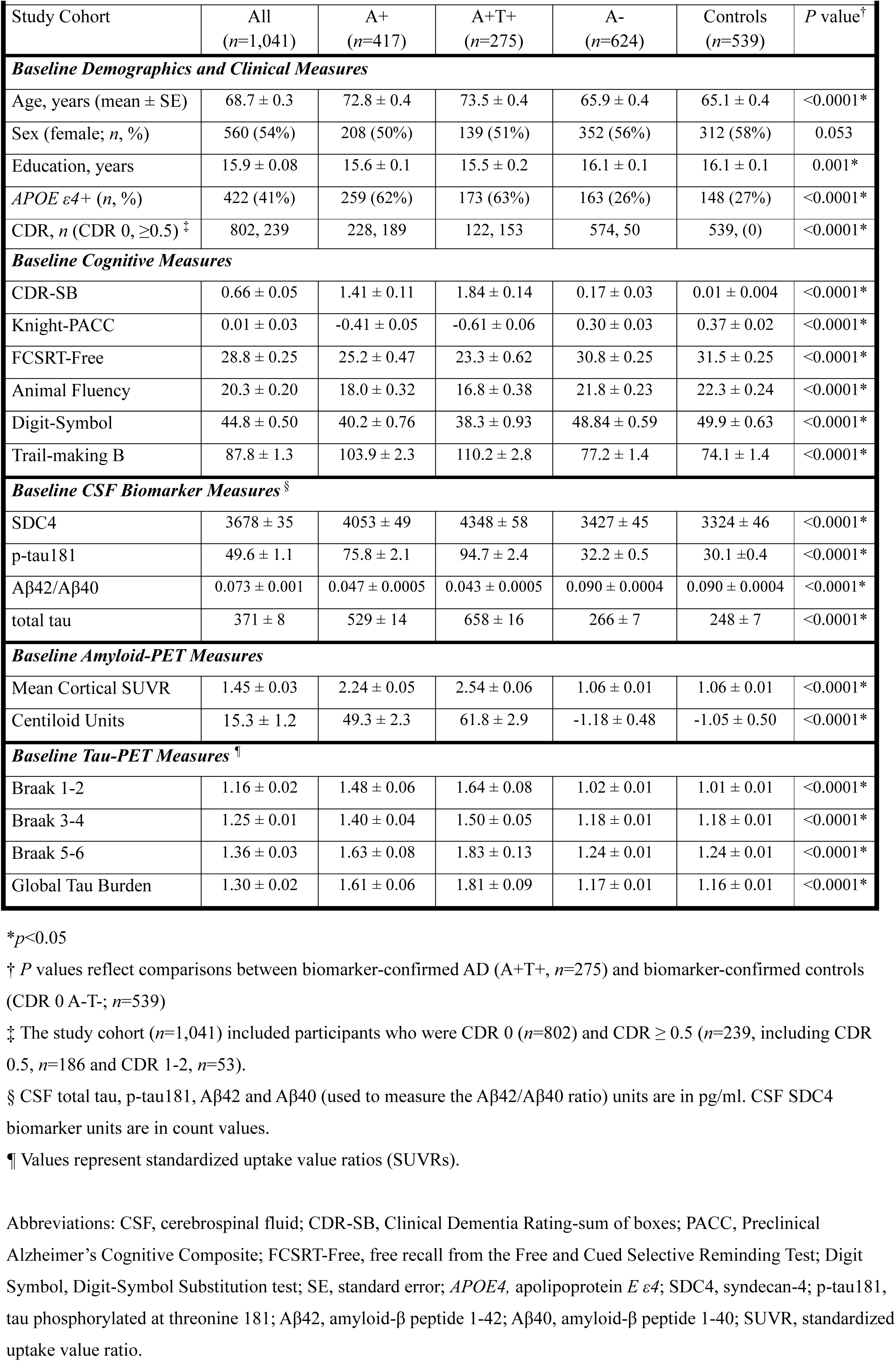
Demographic, Genotypic, Cognitive, CSF and Imaging Biomarker Characteristics of the Study Cohorts.

As expected, CSF levels of established AD biomarkers (total tau, p-tau181, VILIP-1, NFL, Ng, SNAP-25, SYT-1, GFAP, YKL-40, and sTREM2) were significantly higher, and CSF Aβ42/Aβ40 levels were significantly lower, in individuals with biomarker-confirmed AD (A+T+; *n*=275, *p*<0.0001) compared to cognitively unimpaired biomarker-confirmed healthy controls (CDR 0 A-T-, *n*=539) **(Supplementary Table 1)**. CSF levels of established AD markers (total tau, p-tau181, VILIP-1, NFL, Ng, SNAP-25, SYT-1, GFAP, YKL-40 [*p*<0.0001] and sTREM2 [*p*=0.0005]) were significantly higher, and CSF Aβ42/Aβ40 levels were significantly lower (*p*<0.0001), in individuals with biomarker-confirmed preclinical AD (CDR 0 A+T+, *n*=122), compared to biomarker-confirmed healthy controls (CDR 0 A-T-, *n*=539). Mean (± standard error [SE]) of CSF biomarker levels were 605 ± 21 and 248 ± 7 for total tau, 86.8 ± 3.3 and 30.1 ± 0.4 for p-tau181, 0.043 ± 0.0007 and 0.090 ±0.0004 for Aβ42/Aβ40, 3245 ± 98 and 1955 ± 36 for VILIP-1, 6346 ± 263 and 3844 ± 121 for NFL, 4928± 141 and 4281 ± 58 for Ng, 2469 ± 98 and 1671 ± 37 for SNAP-25, 147481 ± 2891 and 108408 ± 1371 for SYT-1, 77386 ± 4321 and 40868 ± 1211 for GFAP, 956325 ± 29587 and 724831 ± 13076 for YKL-40, and 94159 ± 1155 and 18360 ± 536 for sTREM2, in the CDR 0 A+T+ (i.e., preclinical AD) and biomarker-confirmed healthy controls, respectively.

### 3.2. CSF SDC4 levels are increased in preclinical AD

We examined whether CSF SDC4 levels were differentially expressed in the AT sub-cohorts compared to biomarker-confirmed healthy controls (CDR 0 A-T-, *n*=539). CSF SDC4 levels were elevated in the A+T-cohort (mean ± SE, 3483 ± 72, *n*=142) compared to biomarker-confirmed controls (3324 ± 46; *p*=0.03) and were further elevated in the A+T+ cohort (4348 ± 58, *n*=275) compared to A+T- and biomarker-confirmed controls (*p*<0.0001, adjusting for age and sex) (**Fig 1A**). CSF SDC4 levels were also significantly increased in the A+T-group (3483 ± 72, *n*=142) compared to the A-T-group (3362 ±45, *n*=584) (*p*=0.02) (**Fig 1B**). In the preclinical AD cohort, CSF SDC4 levels were higher in the CDR 0 A+T-(3413 ± 74, *n*=106; *p*=0.04) cohort compared to healthy controls (3324 ± 46; *n*=539) and were further increased in the CDR 0 A+T+ cohort (4274 *±* 87, *n*=122) compared to CDR 0 A+T- and controls (*p*<0.0001, adjusting for age and sex).

**Figure 1.**
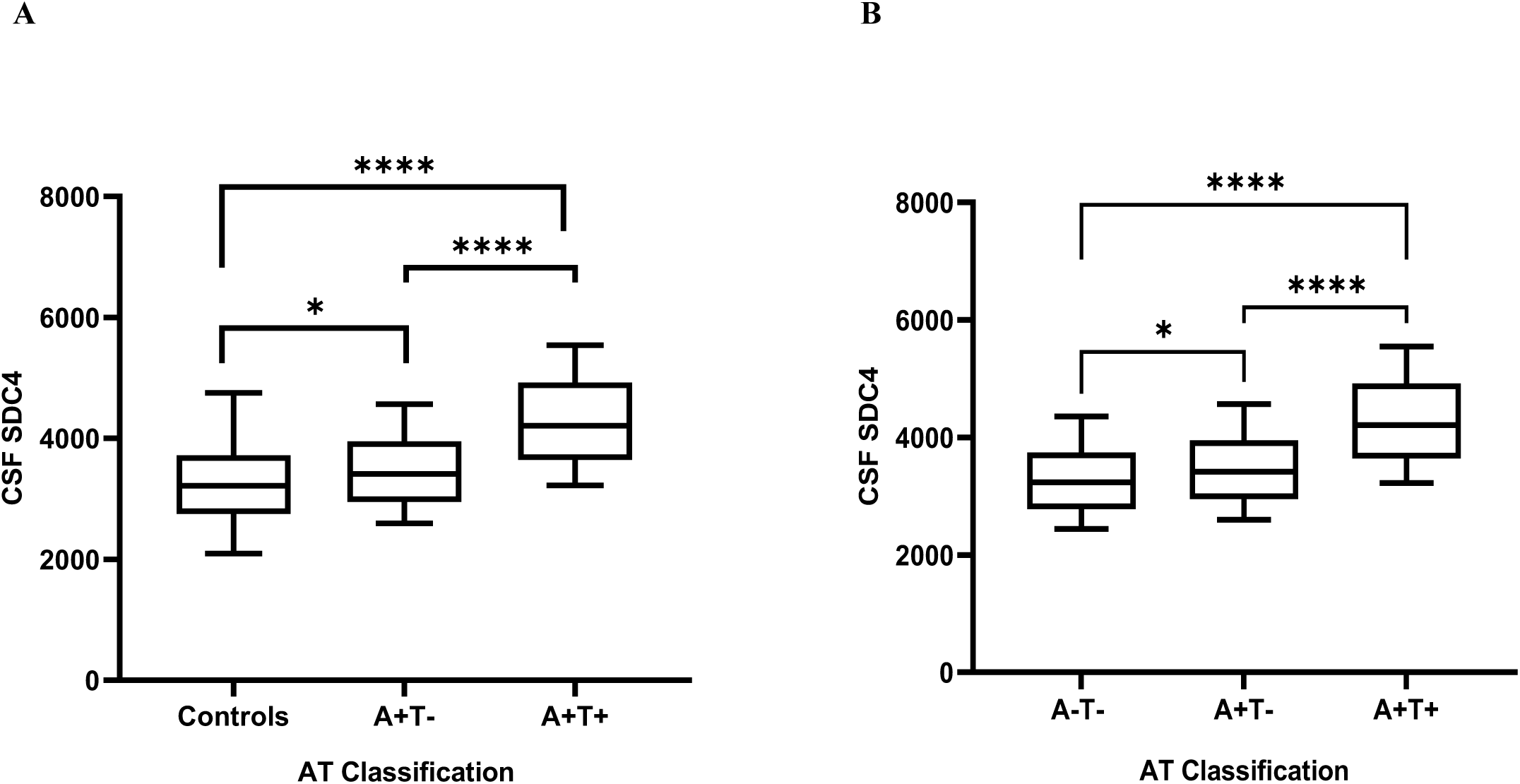
CSF SDC4 levels are elevated in AT subgroups compared to controls. **(A)** CSF SDC4 levels were significantly increased in the A+T-group (mean ± standard error [SE], 3483 ± 72, *n*=142) compared to cognitively unimpaired biomarker-confirmed healthy controls (CDR 0 A-T-, *n*=539) (3324 ± 46, *p*=0.03) and were further significantly elevated in A+T+ (4348 ± 58, *n*=275) compared to A+T- and controls (*p*<0.0001, adjusting for age and sex) in the combined (CDR 0 and CDR ≥0.5) cohort. **(B)** CSF SDC4 levels were significantly increased in the A+T-group (mean ± SE, 3483 ± 72, *n*=142) compared to the A-T-group (3362 ±45, *n*=584) (*p*=0.02). CSF SDC4 levels in the A+T+ group (4348 ± 58, *n*=275) were significantly higher than those in the A+T- and A-T-groups (*p*<0.0001) in the combined (CDR 0 and CDR ≥0.5) cohort. Analyses were adjusted for age and sex. The A-T-cohort (*n*=584) included mostly cognitively unimpaired biomarker confirmed controls (CDR 0 A-T-, *n*=539) and cognitively impaired individuals with no biomarker evidence of AD (CDR ≥ 0.5 A-T-, *n*=45; non-AD). * *p*<0.05, **** *p*<0.0001.

### 3.3. CSF SDC4 levels correlate with fluid and imaging markers of AD pathology

#### 3.3.1. CSF SDC4 levels correlate with other CSF markers of AD pathology

Significant associations between CSF SDC4 and CSF p-tau181 (*r*=0.43), Aβ42/Aβ40 (*r*=-0.26), total tau (*r*=0.43) (*n*=1,032), VILIP-1 (*r*=0.52), NFL (*r*=0.46), Ng (*r*=0.49), SNAP-25 (*r*=0.39), SYT-1 (*r*=0.52), GFAP (*r*=0.50), YKL-40 (*r*=0.69), and sTREM2 (*r*=0.47) (*p*<0.0001) levels were observed in the combined cohort (CDR 0 and CDR ≥ 0.5; *n*=1,041). CSF SDC4 levels also correlated with CSF p-tau181 (*r*=0.40), Aβ42/Aβ40 (*r*=-0.17), total tau (*r*=0.38) (*n*=796, *p*<0.0001), VILIP-1 (*r*=0.49), NFL (*r*=0.38), Ng (*r*=0.49), SNAP-25 (*r*=0.35), SYT-1 (*r*=0.51), GFAP (*r*=0.45), YKL-40 (*r*=0.66), and sTREM2 (*r*=0.52) levels (*p*<0.0001) in the CDR 0 cohort (*n*=802).

To determine whether CSF SDC4 levels were associated with AD pathology across biological disease stages, correlations of CSF SDC4 with other CSF biomarkers were examined in each of the A+ and A+T+ subgroups within the combined (CDR 0 and CDR ≥0.5) and CDR 0 cohorts. CSF SDC4 levels remained closely associated with other CSF AD biomarkers in each of the subgroups as shown in **Supplementary Table 2 and Figure 2**.

**Figure 2.**
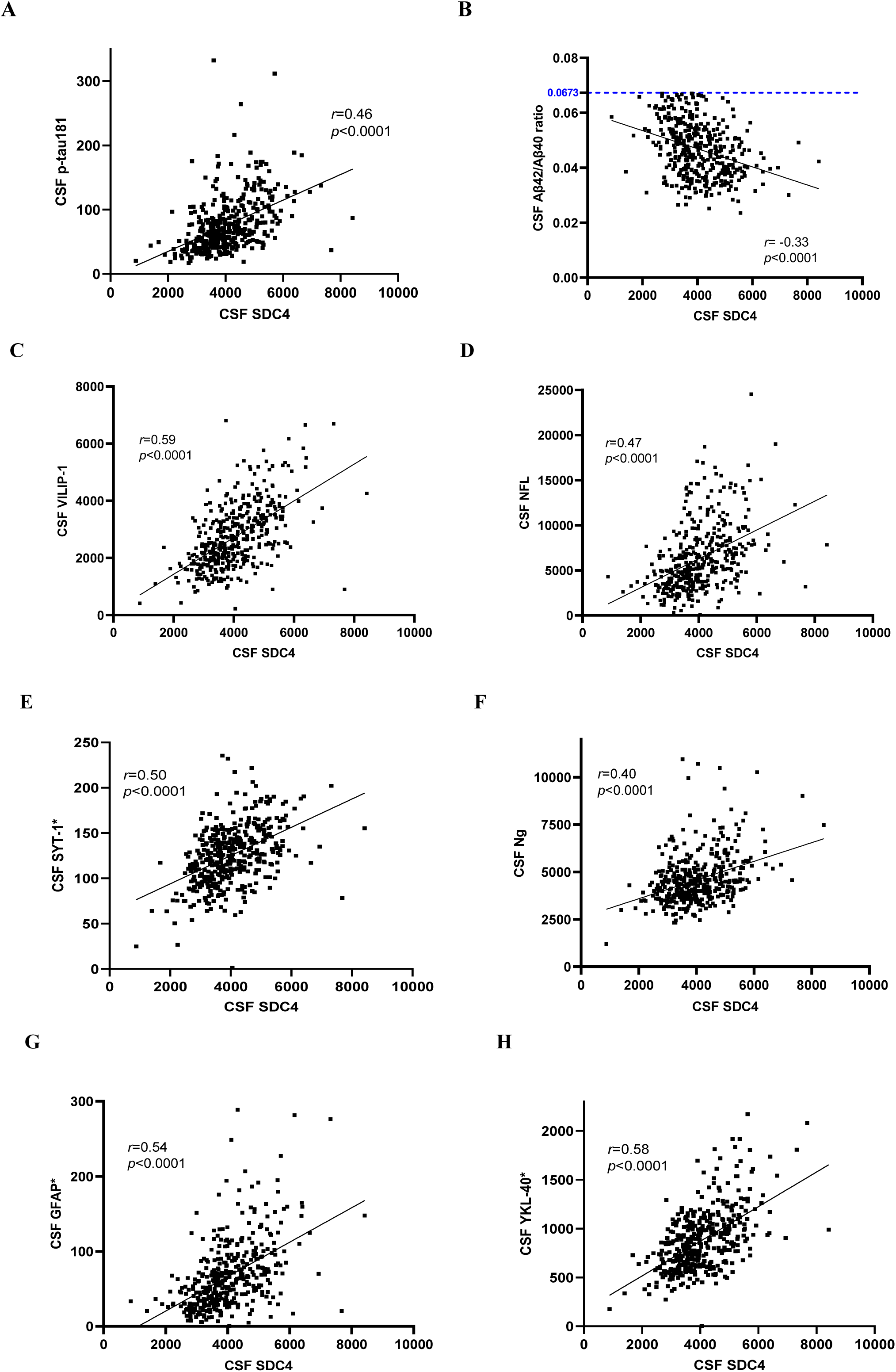
CSF SDC4 Correlations with other CSF Markers of AD Pathology. Significant correlations of CSF SDC4 levels with **(A)** CSF p-tau181 (*r*=0.46) and **(B)** CSF Aβ42/Aβ40 (*r*=-0.33) levels were observed in the Aβ+ cohort (*p*<0.0001, *n*=412). CSF SDC4 levels were also closely associated with CSF levels of neuronal injury markers including **(C)** VILIP-1 (*r*=0.59) and **(D)** NFL (*r*=0.47), synaptic injury markers including **(E)** SYT-1 (*r*=0.50) and **(F)** Ng (*r*=0.40), and markers of astrocytic reactivity including **(G)** GFAP (*r*=0.54), and **(H)** YKL-40 (*r*=0.58) in the Aβ+ cohort (*n*=417, *p*<0.0001). *Values were transformed by dividing the original values by 1000. CSF p-tau181, total tau, Aβ42 and Aβ40 levels were measured in pg/ml. All other CSF biomarker values were measured using count values from the Explore HT Olink proteomics panel. CSF Aβ-positivity was determined using the CSF Aβ42/Aβ40 cut-off of <0.0673 as shown in **Fig 2B**.

#### 3.3.2. CSF SDC4 levels correlate with global and regional tau-PET burden

We investigated the relationship between CSF SDC4 levels and tau pathology using tau-PET estimates of global and regional tau burden in the subset of participants who had at least one tau-PET scan (*n*=302), adjusting for age and sex. In this cohort, CSF SDC4 levels were closely associated with global (*r*=0.25), Braak 1-2 (*r*=0.20), Braak 3-4 (*r*=0.19), and Braak 5-6 (*r*=0.22) estimates of tau burden (*p*<0.0001). When only the CDR 0-0.5 (*n*=293) subset of this cohort was examined, CSF SDC4 levels also correlated with global tau (*r*=0.25, *p*<0.0001), Braak 1-2 (*r*=0.17, *p*=0.004), Braak 3-4 (*r*=0.18, *p*=0.002), and Braak 5-6 (*r*=0.25, *p*<0.0001) estimates. Similar results were obtained when only tau-PET positive participants (i.e., whose global tau burden ≥ 1.22) were examined (*n*=135); CSF SDC4 levels were closely associated with global tau burden (*r*=0.34, *p*<0.0001), Braak 1-2 (*r*=0.23, *p*=0.009), Braak 3-4 (*r*=0.20, *p*=0.02), and Braak 5-6 (*r*=0.28, *p*<0.0001) estimates. **Supplementary Table 3** summarizes CSF SDC4 correlations with global and Braak stage estimates of tau burden in each of the study sub-cohorts.

Notably, CSF SDC4 correlated with regional tau burden in regions known to be involved in early AD, including the entorhinal cortex (*r*=0.20), fusiform (*r*=0.21), inferior temporal (*r*=0.24), middle temporal (*r*=0.25), and temporal pole (*r*=0.20) (*p*<0.0001, *n*=302). Similar results were obtained when only tau-PET positive participants were examined (*n*=135) **(Supplementary Table 4)**.

#### 3.3.3. CSF SDC4 levels correlate with global and regional amyloid-PET burden

CSF SDC4 levels correlated with global amyloid burden measured using the mean cortical SUVR (*r*=0.17, *p*<0.0001) or Centiloid Units (*r*=0.18, *p*<0.0001) in the subset of participants who had amyloid-PET scans (*n*=719) adjusting for age, sex, and *APOE* ε*4*. CSF SDC4 levels significantly correlated with regional amyloid-PET in almost all examined brain regions in the (larger) subset of participants who had amyloid-PET using PiB (*n*=528) **(Supplementary Table 5)**.

### 3.4. Higher CSF SDC4 levels are associated with worse baseline cognition

Higher CSF SDC4 levels were associated with worse cognition as measured by higher CDR-SB (*r*=0.15, *n*=1,041) and lower Knight-PACC (*r*=-0.18), episodic memory (FCSRT-Free, *r*=-0.15), and language (Animal Fluency, *r*=-0.13) (*n*=1,018) scores in the combined cohort (*p*<0.0001), adjusting for age, sex, education, and *APOE* ε*4*. Similar associations were observed in the Aβ+ cohort; CSF SDC4 levels were associated with higher CDR-SB (*r*=0.13, *p*=0.007, *n*=417) and lower Knight-PACC (*r*=-0.20, *p*<0.0001), FCSRT-Free (*r*=-0.13, *p*=0.01), and Animal Fluency (*r*=-0.20, *p*<0.0001) (*n*=408) scores, adjusting for age, sex, education, and *APOE* ε*4* (**Figure 3**).

**Figure 3.**
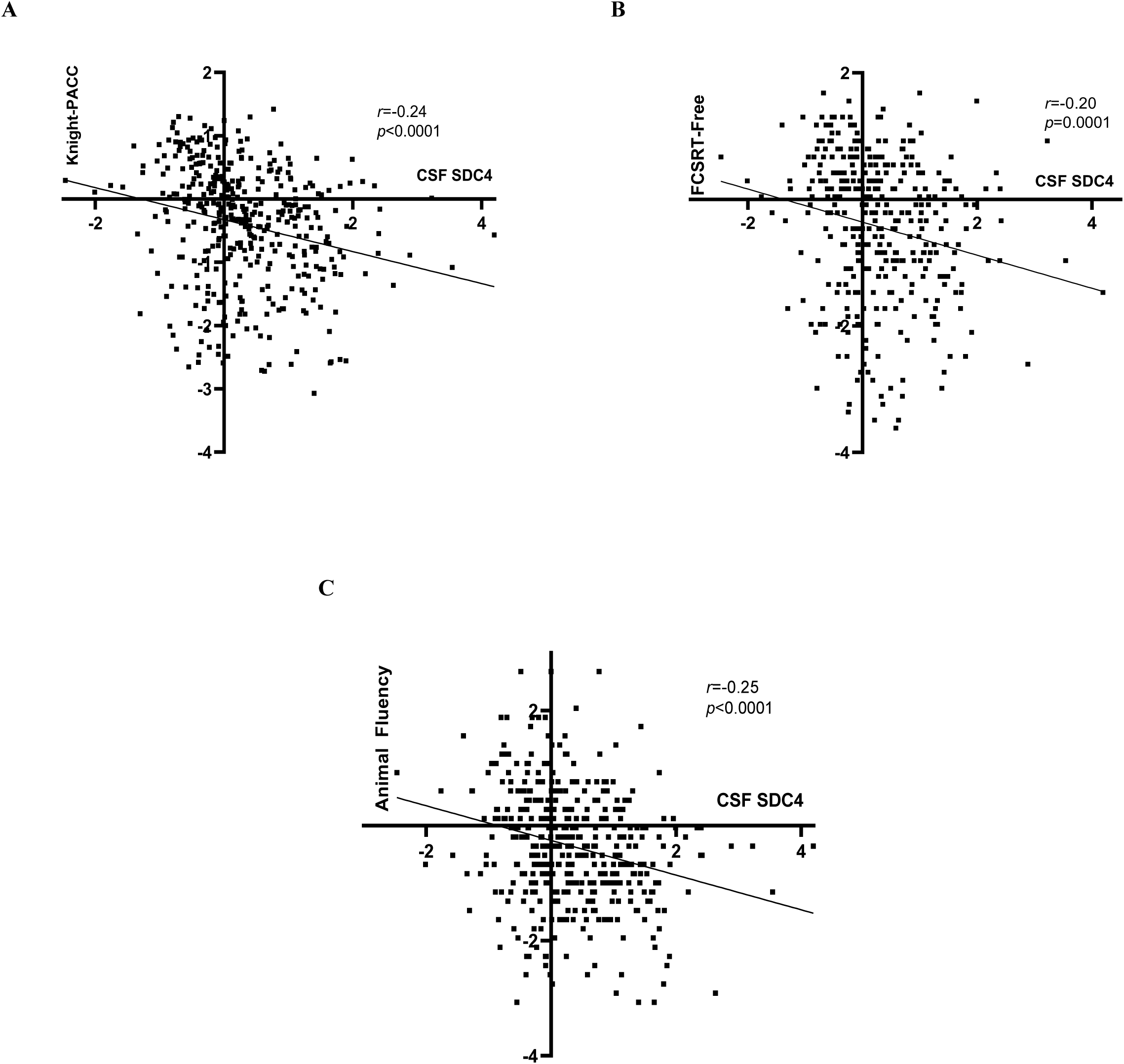
CSF SDC4 Correlations with Cognitive Outcomes. Unadjusted correlation coefficients for CSF SDC4 levels with cognitive outcomes in the Aβ+ cohort are shown. Higher baseline CSF SDC4 levels were associated with worse cognition, including lower **(A)** Knight-PACC scores (unadjusted *r*=-0.24, *p*<0.0001; adjusted *r*=-0.20, *p*<0.0001) **(B)** FCSRT-Free (episodic memory; unadjusted *r*=-0.20, *p*=0.0001; adjusted *r*=-0.13, *p*=0.01), and **(C)** Animal Fluency (language; unadjusted *r*=-0.25, *p*<0.0001; adjusted *r*=-0.20, *p*<0.0001) scores in the Aβ+ (*n*=408) cohort. Cognitive measures and CSF SDC4 levels were standardized to *z* scores.

Baseline CSF SDC4 levels also correlated with lower cognitive scores in each of the CDR 0-0.5 (*n*=988), Aβ+ CDR 0-0.5 (*n*=367), CDR 0 (*n*=802) and Aβ+ CDR 0 (*n*=228) cohorts. No significant associations were observed between CSF SDC4 and baseline executive function measures (i.e., Digit-Symbol or Trails-B) in any of the cohorts after adjusting for covariates. Adjusted correlation coefficients of CSF SDC4 with cognition in the study cohorts are shown in **Supplementary Table 6**.

#### 3.4.1. CSF SDC4 correlations with cognition are mainly mediated by tau

Due to the close associations of CSF SDC4 with CSF or imaging markers of tau pathology in our cohort, we examined whether the global tau burden, which reflects the overall severity of brain tau pathology, mediated the association between CSF SDC4 and cognitive outcomes. CSF SDC4, amyloid-PET, tau-PET, and cognitive scores were all standardized to *z* scores prior to analysis. CSF SDC4 levels were closely associated with CDR-SB (*r*=0.15), Knight-PACC (*r*=-0.18), FCSRT-Free (*r*=-0.15), and Animal Fluency (*r*=-0.14) scores (*p*<0.0001) in the subset of participants who had at least one tau-PET scan (*n*=302), adjusting for age, sex, education, and *APOE* ε*4*. The global tau burden fully mediated the relationship between CSF SDC4 and CDR-SB (β estimate of indirect effect ± SE, 95% confidence interval [CI]; 0.182 ± 0.07, 0.075 to 0.342), Knight-PACC (−0.125 ± 0.048, −0.236 to −0.053), FCSRT-Free (−0.075 ± 0.034, −0.152 to −0.021), and Animal Fluency (−0.108 ± 0.045, −0.216 to −0.040) scores, adjusting for age, sex, education, and *APOE* ε*4.* Similarly, the global tau burden fully mediated associations of CSF SDC4 (β estimate ± SE, 95% CI) with CDR-SB (0.072 ± 0.030, 0.023 to 0.143), Knight-PACC (−0.063 ± 0.025, −0.121 to −0.024), FCSRT-Free (−0.056 ± 0.030, −0.126 to −0.011), and Animal Fluency (−0.057 ± 0.026, −0.118 to −0.017) scores in the CDR 0−0.5 subset of participants who had tau-PET scans (*n*=293).

For comparison, the global amyloid burden, measured using the mean cortical SUVR, also mediated CSF SDC4 associations with CDR-SB (β ± SE, 95% CI; 0.051 ± 0.016, 0.025 to 0.088; *n*=719) and Knight-PACC (−0.035 ± 0.011, −0.060 to −0.017) in all participants, and the CDR 0−0.5 subset of participants (CDR-SB, 0.024 ± 0.008, 0.011 to 0.042; Knight-PACC, −0.020 ± 0.007, −0.037 to −0.009; *n*=706), who had amyloid-PET scans; however, the global tau burden was a stronger mediator of CSF SDC4 associations with CDR-SB and Knight-PACC in this cohort. The global amyloid burden only partially mediated the relationship between CSF SDC4 and FCSRT-Free (−0.024 ± 0.010, −0.047 to −0.008) or Animal Fluency (−0.031 ± 0.011, −0.057 to −0.013) scores in the subset of participants who had amyloid-PET scans (*n*=719; adjusting for age, sex, education, and *APOE* ε*4*). Similarly, the global amyloid burden partially mediated the relationship between CSF SDC4 and FCSRT-Free (−0.018 ± 0.008, −0.038 to −0.005) or Animal Fluency (−0.018 ± 0.008, −0.036 to −0.006) scores in the CDR 0−0.5 subset of participants who had amyloid-PET scans (*n*=706).

Importantly, in the combined cohort (*n*=302), the global tau burden remained a significant mediator of CSF SDC4 effects on CDR-SB (0.085 ± 0.032, 0.032 to 0.157), Knight-PACC (−0.064 ± 0.024, −0.122 to −0.024), FCSRT-Free (−0.044 ± 0.025, −0.100 to −0.003), and Animal Fluency (−0.047 ± 0.024, −0.105 to −0.013) scores after adjusting for the mediating effects of global amyloid burden (and the other covariates).

### 3.5. Higher baseline CSF SDC4 levels predict more rapid cognitive decline

Cognitive outcomes worsened at an average annual rate (± SE) of 0.195 ± 0.014 and 0.135 ± 0.009 for CDR-SB, −0.058 ± 0.003 for Knight-PACC, −0.282 ± 0.032 and −0.036 ± 0.004 for FCSRT-Free, −0.311 ± 0.017 and −0.051 ± 0.003 for Animal Fluency, −0.817 ± 0.056 and −0.063 ± 0.004 for Digit-Symbol, and 2.33 ± 0.143 and 0.058 ± 0.004 for Trails-B, for untransformed and standardized scores, respectively, over follow-up in the combined cohort, adjusting for age, sex, education, and *APOE* ε*4* (*p*<0.0001). Linear mixed models examined whether baseline CSF SDC4 levels predicted annual rates of cognitive decline adjusting for age, sex, education and *APOE* ε*4*. In the subset of participants who had at least one follow-up cognitive assessment (*n*=898), higher standardized baseline CSF SDC4 levels, examined as a continuous variable, were associated with more rapid decline in global cognition (β estimate ± SE, CDR-SB, 0.15 ± 0.02 and 0.10 ± 0.012 and Knight-PACC, −0.025 ± 0.004, *p*<0.0001) over follow-up. Standardized baseline CSF SDC4 levels also predicted more rapid worsening in episodic memory (FCSRT-Free, −0.20 ± 0.04 and −0.024 ± 0.005, *p*<0.0001), language (Animal Fluency, −0.083 ± 0.02 and −0.014 ± 0.004, *p*=0.0004), and executive functions (Digit-Symbol, −0.32 ±0.07 and −0.025 ±0.006, *p*<0.0001 and Trails-B, 1.00 ± 0.20 and 0.024 ± 0.005, *p*<0.0001) (for untransformed and standardized cognitive scores, respectively) over time.

In the Aβ+ subset (*n*=340) of this longitudinal cohort, cognitive outcomes worsened at an average annual rate (± SE) of 0.446 ± 0.032 and 0.308 ±0.022 for CDR-SB, −0.107 ± 0.006 for Knight-PACC, −1.00 ±0.075 and −0.126 ± 0.009 for FCSRT-Free, −0.562 ± 0.040 and −0.092 ± 0.007 for Animal Fluency, −1.42 ± 0.11 and −0.110 ± 0.009 for Digit-Symbol, 4.76 ± 0.32 and 0.118 ± 0.008 for Trails-B (for untransformed and standardized scores, respectively) over follow-up, adjusting for age, sex, education, and *APOE* ε*4* (*p*<0.0001). Standardized baseline CSF SDC4 levels predicted rates of progression in global cognition (β estimate ± SE, CDR-SB, 0.21 ± 0.04 and 0.14 ± 0.03, *p*<0.0001 and Knight-PACC, −0.03 ± 0.009, *p*=0.0005), FCSRT-Free (β estimate ± SE, −0.34 ± 0.10 and −0.042 ± 0.013, *p*=0.0014), Animal Fluency (−0.18 ± 0.05 and −0.030 ± 0.009, *p*=0.001), Digit-Symbol (−0.37 ± 0.14 and −0.028 ± 0.011, *p*=0.012), and Trails-B (1.60 ± 0.45 and 0.039 ± 0.011, *p*=0.0005) (for untransformed and standardized scores, respectively) over time in the Aβ+ longitudinal cohort.

To illustrate the predictive ability of CSF SDC4 for cognitive decline in our cohort, analyses were performed examining the ability of baseline CSF SDC4 as a categorical variable (i.e., divided into quartiles) in predicting longitudinal rates of cognitive decline over time, adjusting for age, sex, education, and *APOE* ε*4*. In the combined cohort, CSF SDC4 levels in the upper quartiles were associated with more rapid cognitive decline in CDR-SB (*p*<0.0001) (**Figure 4A**), Knight-PACC (*p*<0.0001), FCSRT-Free (*p*<0.0001), Animal Fluency (*p*=0.0004), Digit-Symbol (*p*=0.0002), and Trails-B (*p*<0.0001) scores over time compared to those in the lower quartiles. Similarly, CSF SDC4 levels in the upper quartiles were associated with more rapid cognitive decline in CDR-SB (*p*=0.0003), Knight-PACC (*p*=0.018), FCSRT-Free (*p*=0.01) (**Figure 4B-D**), Animal Fluency (*p*=0.04), Digit-Symbol (*p*=0.04), and Trails-B (*p*=0.04) scores compared to those in the lower quartiles in the longitudinal Aβ+ cohort (*n*=340). Adjusted annual rates of cognitive decline in each of the CSF SDC4 quartiles in the longitudinal combined and Aβ+ cohorts are summarized in **Table 2**.

**Figure 4.**
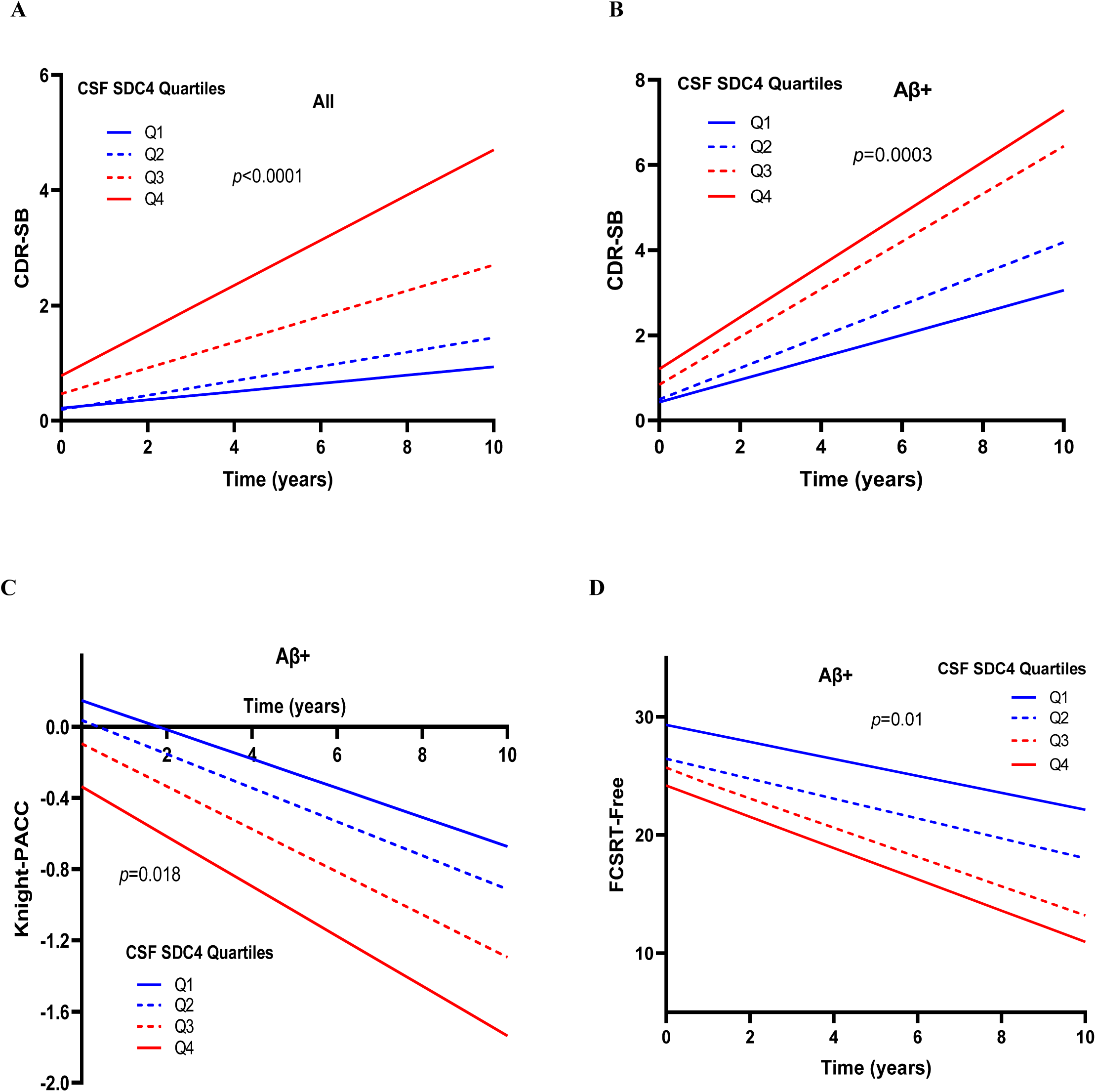
Adjusted annual rates of cognitive decline as a function of CSF SDC4 levels. **(A)** Participants (*n*=898) with longitudinal cognitive assessments whose baseline CSF SDC4 levels were in the upper quartiles demonstrated more rapid progression in CDR-SB scores over time compared to those whose SDC4 levels were in the lower quartiles (*p*<0.0001). Cut-off values for CSF SDC4 levels in the combined longitudinal cohort were: 2975 (25^th^ percentile), 3571 (median, 50^th^ percentile), and 4277 (75^th^ percentile). **(B-D)** Aβ+ participants who had longitudinal cognitive assessments (*n*=340) whose baseline CSF SDC4 levels were in the upper quartiles had more rapid cognitive decline compared to those whose CSF SDC4 levels were in the lower quartiles as demonstrated by a more rapid increase in **(B)** CDR-SB scores (*p*=0.0003) and a more rapid decline in **(C)** Knight-PACC (*p*=0.018) and **(D)** FCSRT-Free (*p*=0.01) scores over time. Analyses used untransformed cognitive scores and were adjusted for age, sex, education and *APOE* ε*4*. Cut-off values for CSF SDC4 levels in the Aβ+ longitudinal cohort were: 3391 (25^th^ percentile), 3959 (median, 50^th^ percentile), and 4694 (75^th^ percentile). Adjusted annual rates of cognitive decline for each of the CSF SDC4 quartiles are shown in **Table 2**. Shown intercepts are model estimates adjusting for age, sex, education, and *APOE* ε*4*.

**Table 2.** Adjusted Annual Rates of Cognitive Decline as a Function of CSF SDC4 Quartiles^†^.

|  |  |  |  |  |  |  |
| --- | --- | --- | --- | --- | --- | --- |
| **Adjusted Estimated Rates of Change in Cognitive Outcomes in all participants (*n*=898)<sup>‡</sup>** |  |  |  |  |  |  |
| **SDC4 Quartile** | **CDR-SB** | **Knight-PACC** | **FCSRT-Free** | **Animal Fluency** | **Digit-Symbol** | **Trails-B** |
| Q1 | 0.072 ± 0.026 | -0.036 ± 0.005 | -0.109 ± 0.058 | -0.230 ± 0.032 | -0.605 ± 0.121 | 1.587 ± 0.256 |
| Q2 | 0.125 ± 0.025 | -0.049 ± 0.005 | -0.233 ± 0.058 | -0.280 ± 0.032 | -0.613 ± 0.103 | 1.985 ± 0.260 |
| Q3 | 0.224 ± 0.026 | -0.068 ± 0.006 | -0.349 ± 0.064 | -0.372 ± 0.037 | -0.847 ± 0.107 | 2.700 ± 0.292 |
| Q4 | 0.392 ± 0.029 | -0.094 ± 0.007 | -0.571 ± 0.075 | -0.433 ± 0.043 | -1.233 ± 0.118 | 3.786 ± 0.338 |
| *p* value | <0.0001* | <0.0001* | <0.0001* | 0.0004* | 0.0002* | <0.0001* |
| **Adjusted Estimated Rates of Change in Cognitive Outcomes in Aβ+ participants (*n*=340)<sup>§</sup>** |  |  |  |  |  |  |
| Q1 | 0.263 ± 0.060 | -0.082 ± 0.011 | -0.717 ± 0.133 | -0.453 ± 0.068 | -1.076 ± 0.212 | 3.949 ± 0.591 |
| Q2 | 0.369 ± 0.060 | -0.100 ± 0.012 | -0.842 ± 0.139 | -0.464 ± 0.073 | -1.148 ± 0.212 | 4.150 ± 0.616 |
| Q3 | 0.560 ± 0.062 | -0.121 ± 0.015 | -1.238 ± 0.151 | -0.690 ± 0.081 | -1.720 ± 0.221 | 5.160 ± 0.680 |
| Q4 | 0.608 ± 0.069 | -0.143 ± 0.013 | -1.323 ± 0.176 | -0.708 ± 0.099 | -1.741 ± 0.231 | 6.295 ± 0.757 |
| *p* value | 0.0003* | 0.018* | 0.010* | 0.039* | 0.044* | 0.043* |

\**p*<0.05
<sup>†</sup> Values represent β estimates (± standard error) for the CSF SDC4 quartile\*time interaction which estimate annual rates of change in the corresponding cognitive outcome (using untransformed cognitive scores) for each CSF SDC4 quartile, adjusting for age, sex, education, and *APOE ε4*
<sup>‡</sup> Analyses included *n*=898 participants; CDR 0 *n*=705, CDR ≥0.5 *n*=193. Cut-off values for CSF SDC4 levels were: 2975 (25<sup>th</sup> percentile), 3571 (median, 50<sup>th</sup> percentile), and 4277 (75<sup>th</sup> percentile)
<sup>§</sup> Analyses included *n*=340 participants; CDR 0 *n*=194, CDR ≥0.5 *n*=146. Cut-off values for CSF SDC4 levels were: 3391 (25<sup>th</sup> percentile), 3959 (median, 50<sup>th</sup> percentile), and 4694 (75<sup>th</sup> percentile).

### 3.6. Higher baseline CSF SDC4 levels predict more rapid progression of amyloid pathology

In the subset of participants who had longitudinal amyloid-PET scans (*n*=370), the global amyloid burden progressed at an average annual rate of 0.046 ± 0.003 and 0.084 ± 0.006 for untransformed and standardized mean cortical SUVR, respectively, and 1.31 ± 0.14 for Centiloid Units over follow-up, adjusting for age, sex and *APOE* ε*4* (*p*<0.0001).

To determine whether CSF SDC4 was associated with the progression of brain amyloid pathology, we examined whether baseline CSF SDC4 levels, examined as a continuous variable, predicted rates of progression of global amyloid burden over time. Higher standardized baseline CSF SDC4 levels predicted faster progression in global amyloid burden using the untransformed and standardized measures of mean cortical SUVR (β estimate± SE, 0.009 ± 0.004 and 0.016 ± 0.007, *p*=0.013, respectively) or Centiloid Units (0.36 ± 0.16, *p*=0.022) over follow-up, adjusting for age, sex, and *APOE* ε*4*. To illustrate this, we divided baseline CSF SDC4 levels into terciles using the 33^rd^ (2974) and 67^th^ (3678) percentile values and examined their ability to predict annual rates of progression in global amyloid burden. CSF SDC4 levels in the upper tercile were associated with more rapid progression in mean cortical SUVR (*p*=0.0016) or Centiloid Units (*p*=0.0023) compared to those in the lower two terciles. **Supplementary** Figure 1 demonstrates adjusted annual rates of global amyloid progression over follow-up as a function of CSF SDC4 terciles.

### 3.7. Higher baseline CSF SDC4 levels predict more rapid progression of tau pathology

Tau-PET estimates progressed at an average annual rate of 0.029 ± 0.005 and 0.094 ± 0.018 for global tau, 0.027 ± 0.006 and 0.080 ± 0.018 for Braak 1-2, 0.016 ± 0.004 and 0.080 ± 0.018 for Braak 3-4, 0.027 ± 0.006 and 0.065 ± 0.013 for Braak 5-6 (*p*<0.0001) (for untransformed and standardized tau-PET estimates, respectively) over follow-up, adjusting for age and sex. We examined whether baseline CSF SDC4 levels, examined as a continuous variable, predicted rates of progression in tau-PET estimates over time in the subset of participants who had longitudinal tau-PET scans (*n*=92), adjusting for age and sex. In these analyses, higher standardized baseline SDC4 levels were associated with significantly more rapid progression in global (β estimate ± SE, 0.018 ± 0.005 and 0.057 ± 0.016, *p*=0.0005), Braak 3-4 (0.019 ± 0.003 and 0.091 ± 0.015, *p*<0.0001), and Braak 5-6 (0.016 ± 0.005 and 0.039 ± 0.012, *p*=0.0022), but not Braak 1-2 (0.008 ± 0.006 and 0.024 ± 0.017, *p*=0.15), estimates of tau-PET burden (for untransformed and standardized tau-PET measures, respectively) over follow-up.

To illustrate the predictive ability of CSF SDC4 levels for progression in tau-PET measures over time, we divided baseline CSF SDC4 levels into terciles using the 33rd (2887) and 67^th^ (3823) percentile values. CSF SDC4 levels in the upper tercile were associated with more rapid progression in global tau (*p*=0.027), Braak 3-4 (*p*=0.0026), Braak 5-6 (*p*=0.041), but not Braak1-2 (*p*=0.076), estimates compared to the middle and lower terciles over follow-up, adjusting for age and sex. Progressively higher rates of progression in global, Braak 3-4, and Braak 5-6 tau-PET burden were observed with higher CSF SDC4 terciles as shown in **Figure 5**.

**Figure 5.**
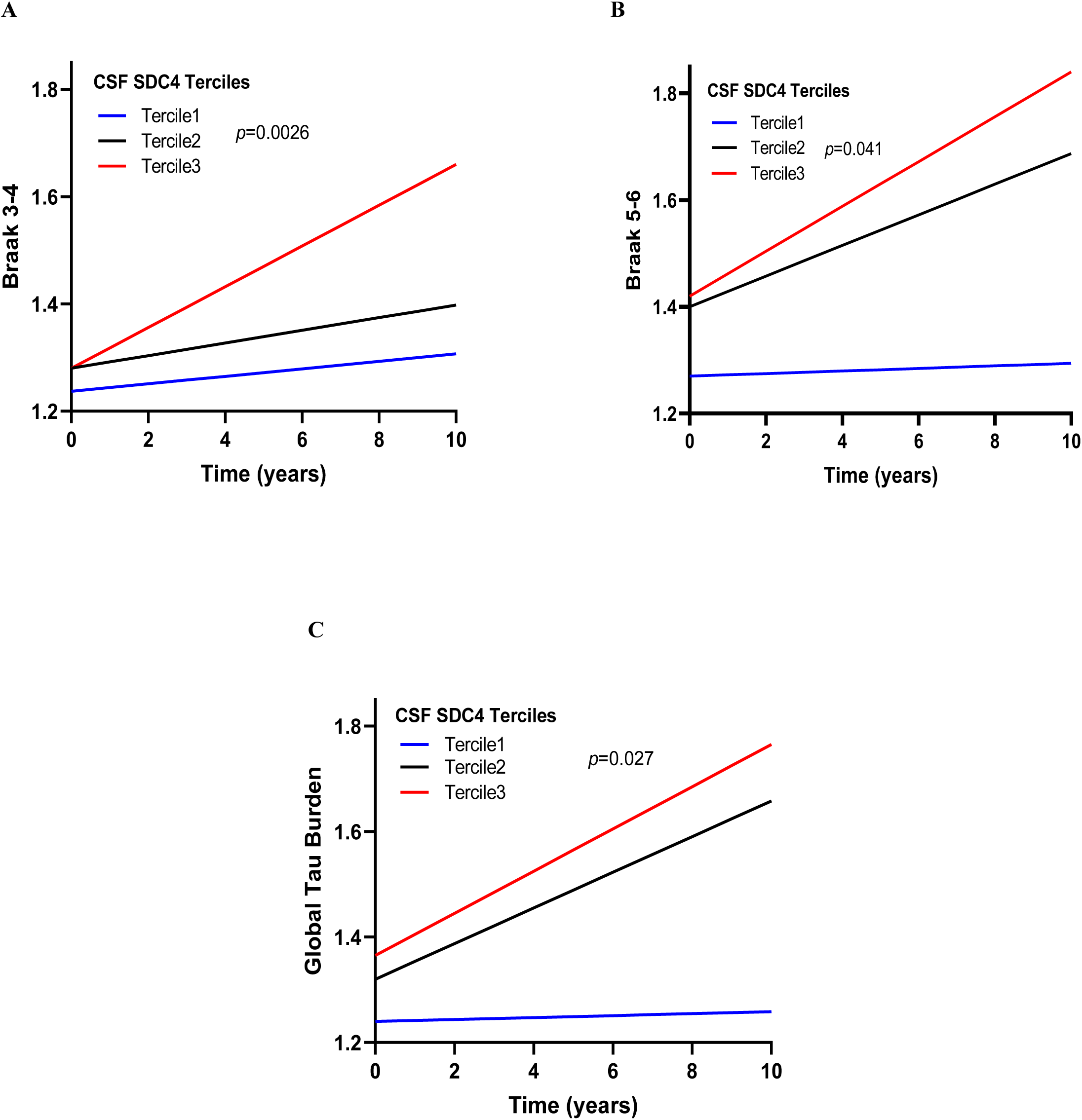
Adjusted annual rates of tau-PET progression as a function of CSF SDC4 levels. Participants (*n*=92) with longitudinal tau-PET assessments whose baseline CSF SDC4 levels were in the upper tercile demonstrated more rapid progression in **(A)** Braak 3-4 (*p*=0.0026), **(B)** Braak 5-6 (*p*=0.041), and **(C)** global estimates (*p*=0.027) of tau-PET burden over time compared to those whose CSF SDC4 levels were in the middle or lower terciles, adjusting for age and sex. Adjusted annual rates of tau-PET progression (using untransformed tau-PET measures) in the lower, middle, and upper CSF SDC4 terciles, respectively, were 0.007 ± 0.006, 0.012 ± 0.005 and 0.038 ± 0.007 for Braak 3-4, 0.002 ± 0.011, 0.029 ± 0.009, 0.042 ± 0.013 for Braak 5-6, and 0.002 ± 0.010, 0.034 ± 0.009, and 0.040 ± 0.013 for global tau-PET (adjusting for age and sex). Cut-off values for CSF SDC4 levels were: 2887 (33^rd^ percentile) and 3823 (67^th^ percentile). Shown intercepts are model estimates adjusting for age and sex.

### 3.8. Inferred trajectories of CSF biomarkers along AD progression

We used trajectory inference methods to estimate and compare the trajectories of CSF SDC4 and established AD biomarkers on a scale which represents a pseudo-time of AD progression. In these analyses, each participant was assigned a point along the pseudo-time, and individual proteins were plotted to demonstrate their trajectories over a granular estimate of disease progression. Our pseudo-time model depicts the sequential progression of participants through the following disease stages: (CDR 0 A-T-), (CDR 0 A+T-), (CDR 0 A+T+), and (CDR ≥ 0.5 A+T+). Spectral embedding from which the pseudo-time variable was estimated is shown (**Figure 6A**).

**Figure 6.**
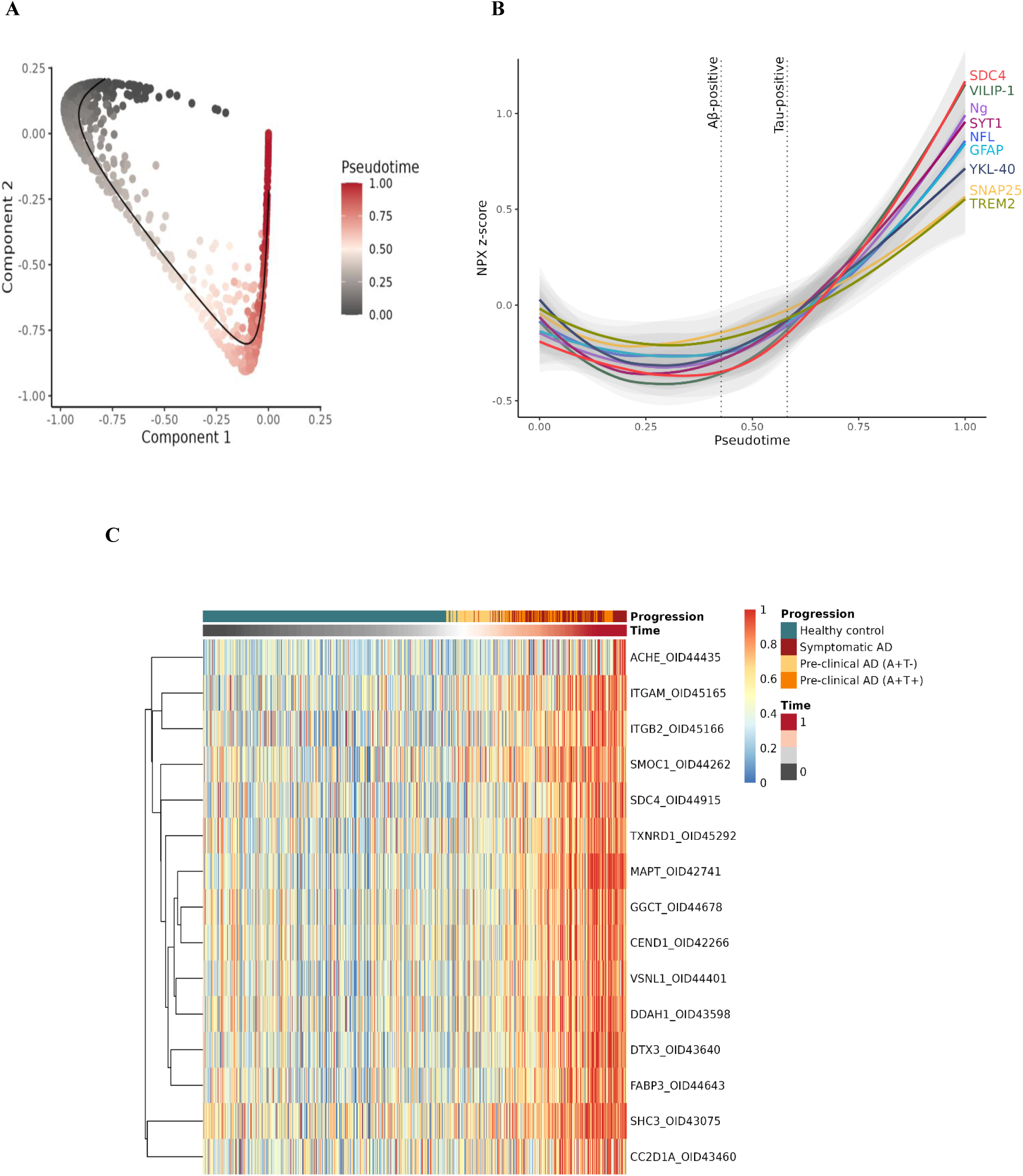
Pseudo-time models of CSF SDC4 and other AD Biomarkers. **(A)** Visualization of the spectral embedding components which were used to infer the trajectory of AD progression along pseudo-time. **(B)** Trajectory inference methods were used to estimate and compare the trajectories of CSF SDC4 and established CSF AD biomarkers (VILIP−1, NFL, Ng, SNAP-25, SYT−1, GFAP, YKL-40, and sTREM2) on a scale which represents a pseudo-time of the progression of AD pathology. The points of Aβ-positivity and tau-positivity are shown as dashed lines. The 95% confidence intervals are shown as gray bands around the trajectory lines for each biomarker. These analyses included NPX (normalized protein expression values reported on a log2 scale) which were adjusted for age, sex, and mean protein levels and standardized to *z* scores prior to analysis. **(C)** Random Forest models estimated the importance of all CSF proteins (*n*=5,416) in the Explore HT Olink panel in predicting the pseudo-time models. SDC4 was among the top 10 proteins (i.e., the other 9 proteins were MAPT, SHC3, SMOC1, DTX3, ITGAM, ITGB2, ACHE, FABP3, and TXNRD1) with the highest importance in predicting these models (FDR-corrected *q* value= 3.70E−06). The top 15 most important proteins in these models are shown (unranked) and include proteins which are known to be associated with AD (e.g., MAPT, VILIP−1 (i.e., VSNL), SMOC1, and ITGB2).^54,74^ The heatmap displays samples (columns) ranked according to their position in the inferred pseudo-time trajectory. The dendrogram (left) shows the hierarchical clustering of the top 15 assays (rows). Abbreviations: MAPT, microtubule-associated protein tau; SHC3, SHC adaptor protein-3; SMOC1, secreted modular calcium-binding protein−1; DTX3, deltex E3 ubiquitin ligase-3; ITGAM, integrin subunit alpha M; ITGB2, integrin beta-2; ACHE, acetylcholinesterase; FABP3, fatty acid binding protein-3; TXNRD1, thioredoxin reductase−1; GGCT, gamma-glutamyl-cyclo-transferase; CEND1, cell cycle exit and neuronal differentiation−1; DDAH1, dimethylarginine dimethyl-amino-hydrolase−1; CC2D1A, coiled-coil and C2 domain containing 1A.

Our models show that elevations in CSF SDC4 levels were observed very early in AD, beginning in close proximity to the point of Aβ-positivity, and increased more robustly following the point of tau-positivity (**Figure 6B**). These findings recapitulate the results of the AT group comparisons in this cohort. Established AD biomarkers (VILIP−1, NFL, Ng, SYT−1, SNAP-25, GFAP, YKL-40, and sTREM2) were also plotted along the pseudo-time (**Figure 6B**). All CSF AD biomarkers increased past the point of Aβ-positivity and continued to increase along the course of AD progression, with possible differences in biomarker trajectories as shown (**Figure 6B**). Of all the AD biomarkers examined in these models, the largest increases in biomarker levels along AD progression were observed in CSF SDC4 followed by CSF VILIP−1 levels.

We then used a random forest model to rank the importance of each of the 5,416 proteins in the Explore HT Olink panel in predicting the ordering of participants along the pseudo-time. CSF SDC4 was among the top 10 proteins with the highest importance in predicting the pseudo-time models (FDR-corrected *q* value=3.70E−06). The top 10 most important proteins also included other proteins known to be associated with AD (e.g., MAPT, SMOC1, and ITGB2)^54^ and others (SHC3, DTX3, ITGAM, ACHE, FABP3, and TXNRD1) (FDR-corrected *q* value=3.70E−06) (**Figure 6C**). Notably, CSF SDC4 demonstrated a higher feature importance (0.44, *q*=3.70E−06) in predicting the pseudo-time models than any of the other established CSF AD biomarkers examined in this cohort: VILIP−1 (0.41, *q*=3.70E−06), SYT−1 (0.28, *q*=3.70E−06), NFL (0.26, *q*=1.07E−05), Ng (0.24, *q*=1.25E−05), GFAP (0.13, *q*=7.77E−04), and YKL-40 (0.09, *q*=0.013). Conversely, SNAP-25 (0.03, *q*=0.96) and sTREM2 (0.03, *q*=0.96) were not significant predictors of the pseudo-time models in this cohort.

### 3.9. Brain endothelial expression of SDC4

Prior studies have consistently demonstrated endothelial expression of SDC4 and the potential value of SDC4 upregulation as a marker of endothelial dysfunction.^16,18,55^ We utilized a published single-cell atlas of the human brain vasculature to confirm the expression of SDC4 in normal brain endothelium.^56^ In normal brains, SDC4 expression is predominantly upregulated in the early stages of endothelial cell development (i.e., from stem cells to endothelial cells) **(Supplementary** Figure 2). Our findings are consistent with prior literature suggesting that SDC4 is abundantly expressed in proliferating endothelium and plays an important role in regulating angiogenesis and other endothelial cell functions.^16,18,55^

SDC4 expression has also been reported in astrocytes.^57^ To examine the possibility of astrocytic contributions to elevated CSF SDC4 levels in our cohort, we compared correlations of CSF SDC4 with CSF levels of established endothelial and astrocytic markers. We found that CSF SDC4 levels correlated more closely with CSF levels of other endothelial markers (i.e., *r*=0.65−0.73; platelet-endothelial cell adhesion molecule [PECAM1] and vascular cell adhesion molecule [VCAM1]) compared to established astrocytic markers (*r*=0.07−0.50; ALDH1L1 and GFAP) in our cohort (*n*=1,041), suggesting a mainly endothelial source for CSF SDC4.

### 3.10. Functional pathway analyses

Functional pathway analyses using STRING^53^ demonstrated interactions between SDC4 and proteins involved in AD pathogenesis (e.g., *APOE*, low-density lipoprotein receptor-related proteins [*LRPs*], and microtubule-associated protein [*MAPT*]) **(Supplementary** Fig 3A). We also demonstrate functional interactions between SDC4 and other established endothelial proteins such as PECAM1 and the vascular endothelial growth factor receptors, FLT4 and KDR **(Supplementary** Fig 3B).

## 4. Discussion

Using CSF proteomics in a large well-characterized longitudinal cohort of AD and healthy controls (*n*=1,041), we here show that CSF SDC4 levels are upregulated in the earliest preclinical stages of AD (i.e., CDR 0 A+T-) and gradually increase as participants progress through preclinical and symptomatic AD. Our pseudo-time models, which infer the trajectories of CSF SDC4 and other AD biomarkers across the clinicopathological continuum of disease progression, suggest that significant CSF SDC4 elevations are already observed by the time of amyloid-positivity, and change more robustly following the point of tau-positivity. Further, CSF SDC4 demonstrated the highest magnitude of elevation over pseudo-time compared to other established AD biomarkers examined in this cohort, including markers of neuronal or synaptic injury and astrocytic or microglial responses. Notably, of more than 5,000 proteins examined in this study, CSF SDC4, but none of the other established CSF AD biomarkers, was among the top 10 proteins with the highest importance in predicting the ordering of participants in the pseudo-time models.

Together, our findings suggest that SDC4 is an early “core” protein in AD pathogenesis whose altered expression begins very early in the disease course and continues with further disease progression, potentially exceeding in magnitude changes observed in other AD-related processes. Further, these data suggest that SDC4, as a promising surrogate of brain endothelial dysfunction, is potentially a stronger predictor of the pseudo-time models, which depict the progression of participants along the clinical and biological stages of disease, than established AD biomarkers that measure other pathologies. Interestingly, transcriptional upregulation of SDC4 in human AD brains has been demonstrated even in areas that are distant from, or devoid of, AD pathology in prior studies, further highlighting its importance as a potential early contributor to AD pathogenesis.^20^

In our cohort, CSF SDC4 levels strongly correlated with fluid and imaging biomarkers of amyloid and tau pathologies across different clinical disease stages, including preclinical AD. CSF SDC4 levels demonstrated close associations with the severity of AD pathology, reflected by higher PET estimates of global and regional amyloid or tau burden in almost all examined brain regions, particularly those known to be involved early in AD (e.g., tau burden in entorhinal, fusiform, inferior and temporal regions).^58^ These findings are consistent with prior reports which suggest SDC4 colocalization with neuritic Aβ plaques and neurofibrillary tangles,^20^ SDC4 associations with vascular tau,^59^ and progressively higher levels of SDC4 expression observed with higher Braak stages.^20^ Notably, our longitudinal analyses suggest that higher baseline SDC4 levels predicted more rapid progression of AD pathology, reflected by significantly faster propagation of both brain Aβ plaques and neurofibrillary tau tangles, over a mean follow-up period of 8 years, further highlighting potential important contributions of SDC4 upregulation to early AD pathogenesis and its close association with predicted trajectories of future disease progression.

Several lines of evidence support an important role for HSPGs, including SDC4, in promoting the aggregation, internalization, and intercellular spread of Aβ and tau.^21,60–64^ HSPGs, including SDC4, are involved in Aβ synthesis, aggregation and fibrillization at the cell surface, impaired clearance, and toxicity.^21,60,65–67^ HSPGs mediate Aβ internalization into the brain endothelium through endocytosis or a functional complex with LRP−1 and *APOE*, thereby regulating amyloid clearance across the brain endothelium.^68^ SDC4 upregulation also promotes tau aggregation into paired helical filaments and intracellular tau uptake.^64^ Soluble pathological tau aggregates are observed within the brain endothelium in AD tauopathy models,^69^ promote intraneuronal tau aggregation,^70^ and interfere with vascular mechanisms of tau clearance.^71^

Importantly, we show-for the first time-that CSF SDC4 levels are closely associated with global and domain-specific cognitive outcomes in AD, and predict future rates of cognitive decline over time, including the earliest preclinical stages, with tau pathology being the primary mediator of the relationship between CSF SDC4 and cognitive outcomes. Higher CSF SDC4 levels correlated with worse baseline cognitive performance, including global cognition, episodic memory, and language functions, even in participants with no or only mild cognitive impairment (i.e., CDR 0−0.5). Higher baseline SDC4 levels reliably predicted worse future cognitive outcomes as demonstrated by more rapid decline in global cognition, episodic memory, language, and executive functions over follow-up. Further, higher CSF SDC4 levels correlated with higher CSF levels of neuronal (tau, NFL, and VILIP−1) and synaptic (Ng, SNAP-25, and SYT−1) injury markers, and CSF markers of astrocytic or microglial responses (YKL-40, GFAP, and sTREM2). Together, these findings suggest important contributions of SDC4 upregulation, and its downstream effects in promoting amyloid and tau pathologies, on synaptic loss and neurodegeneration, which are the main surrogates of cognitive impairment,^72^ and its close association with other pathological substrates of AD.

Interestingly, we show that SDC4 upregulation, which begins very early in AD pathogenesis near the point of amyloid-positivity, predicts future cognitive decline which does not typically begin until a decade or more following significant Aβ deposition. This may be explained by the observation that CSF SDC4 levels continue to increase past amyloid-positivity, and that they reliably predict the progression of tau pathology, a more imminent harbinger of cognitive impairment. This notion is supported by our mediation analyses which suggest that the relationship between CSF SDC4 and cognitive outcomes was mainly mediated by increased tau pathology, and by closer associations of CSF SDC4 with tau, compared to amyloid, pathology in both cross-sectional and longitudinal analyses. Consistent with these findings, CSF SDC4 levels correlated with cognitive outcomes in preclinical AD and with the severity of tau pathology in critical brain regions (e.g., inferior and middle temporal regions, fusiform, and temporal pole) whose tau burden has been shown to predict cognitive outcomes.^73^

Observations from our multidimensional cross-sectional and longitudinal studies, in a large well-characterized AD cohort, support the presence of close associations of CSF SDC4, as a potential surrogate of brain endothelial dysfunction, with the onset, progression, and severity of the two core AD pathologies. To the best of our knowledge, this is the first longitudinal translational study to examine the relationship between CSF SDC4 and cognitive outcomes or other AD biomarkers and estimate the trajectories of CSF SDC4, in relation to established AD markers, over pseudo-time. Our findings support the notion that SDC4 upregulation is an early event in AD pathogenesis which is temporally associated with the onset and progression of amyloid and tau aggregation, correlates with synaptic injury and neurodegeneration, and reliably predicts trajectories of future cognitive decline. Therefore, we propose that SDC4 plays an important role in early AD pathogenesis, and that CSF SDC4 levels offer a useful surrogate to examine brain endothelial contributions to AD progression and cognitive impairment. Our models also suggest that SDC4 upregulation, as a potential proxy to brain endothelial dysfunction, is an early core pathology in AD which can reliably predict the progression of participants across the clinicopathological continuum of AD (i.e., sequentially from CDR 0 A-T- to CDR 0 A+T-to CDR 0 A+T+ to CDR ≥ 0.5 A+T+) to a potentially better extent than markers that measure other AD pathologies (e.g., neuronal/synaptic injury and microglial/astrocytic responses). It will be important for future studies to examine the role of brain endothelial dysfunction in AD pathogenesis and cognitive impairment and to integrate brain endothelial dysfunction into the current biological definitions and predictive disease models of AD.

Strengths of our study include examining a large well-characterized longitudinal AD cohort that has been followed for an average of 8 years with detailed clinical, cognitive, and PET-imaging data, and for whom we have performed in-depth CSF proteomic profiling. Our cohort includes participants across the different clinical and biological disease stages which has allowed us to examine models of biomarker progression across the disease course using pseudo-time. Importantly, as our cohort consists mostly of cognitively normal participants including healthy controls and those in different biological stages of preclinical AD, we were able to extend our pseudo-time models to include participants in the earliest stages of AD pathogenesis near the time of amyloid-positivity. With the availability of a large comprehensive panel of CSF proteins which encompass various biological processes, we demonstrate the importance of CSF SDC4 upregulation in predicting AD progression compared to biomarkers that measure other AD pathologies.

Our study also has a few limitations. As a clinical study, it does not include mechanistic analyses which will be needed to better understand the role of SDC4 in AD pathogenesis and elucidate specific molecular pathways by which SDC4 upregulation contributes to AD progression. Our study does not include longitudinal SDC4 measurements which will be important to evaluate the potential of SDC4 as a dynamic target for novel disease-modifying therapies that reduce or prevent the aggregation or progression of amyloid and tau. While our cohort includes participants of diverse racial/ethnic backgrounds, it predominantly (∼90%) consists of White participants; therefore, it will be important for future studies to validate these findings in larger multiracial cohorts and to examine potential racial differences in SDC4 levels or their associations with cognition and neurodegeneration. Understanding the role of brain endothelial dysfunction in AD and its contribution to cognitive impairment will offer important insights into early disease pathogenesis and help identify novel biomarkers and therapeutic strategies which target early AD pathologies upstream to amyloid and tau.

## Data Availability

Data was obtained from the Knight-ADRC through an approved data request. All data was de-identified. Proteomic data from the Knight ADRC participants can be requested as described: https://knightadrc.wustl.edu/professionals-clinicians/request-center-resources/.

## Acknowledgments

We thank the research volunteers who participated in the studies, from whom these data were obtained, and their supportive families. We thank the Clinical, Fluid Biomarker and Imaging Cores at the Knight ADRC for sample and data collection.

## References

1. Miyakawa T, Shimoji A, Kuramoto R, Higuchi Y. The relationship between senile plaques and cerebral blood vessels in Alzheimer’s disease and senile dementia. Morphological mechanism of senile plaque production. *Virchows Archiv B*, Cell pathology including molecular pathology. Aug 1982;40(2):121–9. doi:10.1007/bf02932857

2. Araki K, Miyakawa T, Katsuragi S. Ultrastructure of senile plaque using thick sections in the brain with Alzheimer’s disease. The Japanese journal of psychiatry and neurology. Mar 1991;45(1):85–9. doi:10.1111/j.1440-1819.1991.tb00510.x

3. Buée L, Hof PR, Bouras C, et al. Pathological alterations of the cerebral microvasculature in Alzheimer’s disease and related dementing disorders. Acta Neuropathol. 1994;87(5):469–80. doi:10.1007/bf00294173

4. Kalaria RN, Hedera P. Differential degeneration of the cerebral microvasculature in Alzheimer’s disease. Neuroreport. Feb 15 1995;6(3):477–80. doi:10.1097/00001756-199502000-00018

5. Kelleher RJ, Soiza RL. Evidence of endothelial dysfunction in the development of Alzheimer’s disease: Is Alzheimer’s a vascular disorder? Am J Cardiovasc Dis. Nov 1 2013;3(4):197–226.

6. Kalaria RN. Cerebral vessels in ageing and Alzheimer’s disease. Pharmacol Ther. 1996;72(3):193–214. doi:10.1016/s0163-7258(96)00116-7

7. Yamazaki Y, Shinohara M, Shinohara M, et al. Selective loss of cortical endothelial tight junction proteins during Alzheimer’s disease progression. Brain. Apr 1 2019;142(4):1077–1092. doi:10.1093/brain/awz011

8. Meyer EP, Ulmann-Schuler A, Staufenbiel M, Krucker T. Altered morphology and 3D architecture of brain vasculature in a mouse model for Alzheimer’s disease. Proc Natl Acad Sci U S A. Mar 4 2008;105(9):3587–92. doi:10.1073/pnas.0709788105

9. Tarawneh R. Microvascular Contributions to Alzheimer Disease Pathogenesis: Is Alzheimer Disease Primarily an Endotheliopathy? Biomolecules. May 13 2023;13(5)doi:10.3390/biom13050830

10. de la Torre JC. Hemodynamic consequences of deformed microvessels in the brain in Alzheimer’s disease. Ann N Y Acad Sci. Sep 26 1997;826:75–91. doi:10.1111/j.1749-6632.1997.tb48462.x

11. Lau S-F, Cao H, Fu AKY, Ip NY. Single-nucleus transcriptome analysis reveals dysregulation of angiogenic endothelial cells and neuroprotective glia in Alzheimer’s disease. Proceedings of the National Academy of Sciences. 2020;117(41):25800. doi:10.1073/pnas.2008762117

12. Yang AC, Vest RT, Kern F, et al. A human brain vascular atlas reveals diverse cell mediators of Alzheimer’s disease risk. *bioRxiv*. 2021:2021.04.26.441262. doi:10.1101/2021.04.26.441262

13. Grammas P. Neurovascular dysfunction, inflammation and endothelial activation: implications for the pathogenesis of Alzheimer’s disease. J Neuroinflammation. Mar 25 2011;8:26. doi:10.1186/1742-2094-8-26

14. Takechi R, Lam V, Brook E, et al. Blood-Brain Barrier Dysfunction Precedes Cognitive Decline and Neurodegeneration in Diabetic Insulin Resistant Mouse Model: An Implication for Causal Link. Front Aging Neurosci. 2017;9:399. doi:10.3389/fnagi.2017.00399

15. Tarawneh R. Biomarkers: Our Path Towards a Cure for Alzheimer Disease. Biomark Insights. 2020;15:1177271920976367. doi:10.1177/1177271920976367

16. Lipphardt M, Dihazi H, Maas J-H, et al. Syndecan-4 as a Marker of Endothelial Dysfunction in Patients with Resistant Hypertension. Journal of Clinical Medicine. 2020;9(9):3051.

17. Elfenbein A, Simons M. Syndecan-4 signaling at a glance. J Cell Sci. Sep 1 2013;126(Pt 17):3799–804. doi:10.1242/jcs.124636

18. Vuong TT, Reine TM, Sudworth A, Jenssen TG, Kolset SO. Syndecan-4 is a major syndecan in primary human endothelial cells in vitro, modulated by inflammatory stimuli and involved in wound healing. J Histochem Cytochem. Apr 2015;63(4):280–92. doi:10.1369/0022155415568995

19. Partovian C, Ju R, Zhuang ZW, Martin KA, Simons M. Syndecan-4 Regulates Subcellular Localization of mTOR Complex2 and Akt Activation in a PKCα-Dependent Manner in Endothelial Cells. Molecular Cell. 2008/10/10/ 2008;32(1):140–149. 10.1016/j.molcel.2008.09.010

20. Lorente-Gea L, García B, Martín C, et al. Heparan Sulfate Proteoglycans Undergo Differential Expression Alterations in Alzheimer Disease Brains. J Neuropathol Exp Neurol. May 1 2020;79(5):474–483. doi:10.1093/jnen/nlaa016

21. Snow AD, Cummings JA, Lake T. The Unifying Hypothesis of Alzheimer’s Disease: Heparan Sulfate Proteoglycans/Glycosaminoglycans Are Key as First Hypothesized Over 30 Years Ago. Review. Frontiers in Aging Neuroscience. 2021-October−04 2021;13doi:10.3389/fnagi.2021.710683

22. Morris JC. The Clinical Dementia Rating (CDR): current version and scoring rules. Neurology. Nov 1993;43(11):2412–4. doi:10.1212/wnl.43.11.2412-a

23. Talbot C, Lendon C, Craddock N, Shears S, Morris JC, Goate A. Protection against Alzheimer’s disease with apoE epsilon 2. Lancet. Jun 4 1994;343(8910):1432–3. doi:10.1016/s0140-6736(94)92557-7

24. Tarawneh R, Lee JM, Ladenson JH, Morris JC, Holtzman DM. CSF VILIP−1 predicts rates of cognitive decline in early Alzheimer disease. Neurology. Mar 6 2012;78(10):709–19. doi:10.1212/WNL.0b013e318248e568

25. Donohue MC, Sperling RA, Salmon DP, et al. The preclinical Alzheimer cognitive composite: measuring amyloid-related decline. JAMA Neurol. Aug 2014;71(8):961–70. doi:10.1001/jamaneurol.2014.803

26. McKay NS, Millar PR, Nicosia J, et al. Pick a PACC: Comparing domain-specific and general cognitive composites in Alzheimer disease research. Neuropsychology. Jul 2024;38(5):443–464. doi:10.1037/neu0000949

27. Grober E, Sanders AE, Hall C, Lipton RB. Free and cued selective reminding identifies very mild dementia in primary care. Alzheimer Dis Assoc Disord. Jul-Sep 2010;24(3):284–90. doi:10.1097/WAD.0b013e3181cfc78b

28. Wechsler D. The psychometric tradition: developing the wechsler adult intelligence scale. Contemporary Educational Psychology. 1981;

29. Armitage SG. An analysis of certain psychological tests used for the evaluation of brain injury. Psychological monographs. 1946;60(1):i.

30. Goodglass H, Kaplan E, Weintraub S. BDAE: The Boston diagnostic aphasia examination. Lippincott Williams & Wilkins Philadelphia, PA; 2001.

31. Barthélemy NR, Saef B, Li Y, et al. CSF tau phosphorylation occupancies at T217 and T205 represent improved biomarkers of amyloid and tau pathology in Alzheimer’s disease. Nature Aging. 2023/04/01 2023;3(4):391–401. doi:10.1038/s43587-023-00380-7

32. Wik L, Nordberg N, Broberg J, et al. Proximity Extension Assay in Combination with Next-Generation Sequencing for High-throughput Proteome-wide Analysis. Mol Cell Proteomics. 2021;20:100168. doi:10.1016/j.mcpro.2021.100168

33. Sun BB, Chiou J, Traylor M, et al. Plasma proteomic associations with genetics and health in the UK Biobank. Nature. 2023/10/01 2023;622(7982):329–338. doi:10.1038/s41586-023-06592-6

34. Jack CR, Jr., Bennett DA, Blennow K, et al. NIA-AA Research Framework: Toward a biological definition of Alzheimer’s disease. Alzheimers Dement. Apr 2018;14(4):535–562. doi:10.1016/j.jalz.2018.02.018

35. Su Y, Blazey TM, Owen CJ, et al. Quantitative Amyloid Imaging in Autosomal Dominant Alzheimer’s Disease: Results from the DIAN Study Group. PLoS One. 2016;11(3):e0152082. doi:10.1371/journal.pone.0152082

36. Su Y, D’Angelo GM, Vlassenko AG, et al. Quantitative analysis of PiB-PET with FreeSurfer ROIs. PLoS One. 2013;8(11):e73377. doi:10.1371/journal.pone.0073377

37. Su Y, Blazey TM, Snyder AZ, et al. Partial volume correction in quantitative amyloid imaging. Neuroimage. 2015;107:55–64.

38. Su Y, Flores S, Hornbeck RC, et al. Utilizing the Centiloid scale in cross-sectional and longitudinal PiB PET studies. NeuroImage: Clinical. 2018/01/01/ 2018;19:406–416. 10.1016/j.nicl.2018.04.022

39. Su Y, Flores S, Wang G, et al. Comparison of Pittsburgh compound B and florbetapir in cross sectional and longitudinal studies. *Alzheimer’s & Dementia: Diagnosis*, Assessment & Disease Monitoring. 2019;11(1):180–190.

40. Chien DT, Bahri S, Szardenings AK, et al. Early clinical PET imaging results with the novel PHF-tau radioligand [F−18]-T807. J Alzheimers Dis. 2013;34(2):457–68. doi:10.3233/jad-122059

41. Mishra S, Gordon BA, Su Y, et al. AV−1451 PET imaging of tau pathology in preclinical Alzheimer disease: Defining a summary measure. Neuroimage. Nov 1 2017;161:171–178. doi:10.1016/j.neuroimage.2017.07.050

42. Braak H, Braak E. Staging of Alzheimer’s disease-related neurofibrillary changes. Neurobiol Aging. May-Jun 1995;16(3):271–8; discussion 278-84. doi:10.1016/0197-4580(95)00021-6

43. Hayes AF. Introduction to mediation, moderation, and conditional process analysis: A regression-based approach. Guilford publications; 2017.

44. Tasaki S, Xu J, Avey DR, et al. Inferring protein expression changes from mRNA in Alzheimer’s dementia using deep neural networks. Nature Communications. 2022/02/03 2022;13(1):655. doi:10.1038/s41467-022-28280-1

45. Csárdi G, Nepusz T. The igraph software package for complex network research. 2006;1695

46. Csárdi G, Nepusz T, Traag V, et al. igraph: Network Analysis and Visualization in R. 2025;doi:doi:10.5281/zenodo.7682609, R package version 2.1.3, https://CRAN.R-project.org/package=igraph

47. Cannoodt R, Saelens W, Sichien D, et al. SCORPIUS improves trajectory inference and identifies novel modules in dendritic cell development. *bioRxiv*. 2016:079509. doi:10.1101/079509

48. Saelens W, Cannoodt R, Todorov H, Saeys Y. A comparison of single-cell trajectory inference methods. Nature Biotechnology. 2019/05/01 2019;37(5):547–554. doi:10.1038/s41587-019-0071-9

49. Schlitzer A, Sivakamasundari V, Chen J, et al. Identification of cDC1- and cDC2-committed DC progenitors reveals early lineage priming at the common DC progenitor stage in the bone marrow. Nature Immunology. 2015/07/01 2015;16(7):718–728. doi:10.1038/ni.3200

50. Wickham H. ggplot2: Elegant Graphics for Data Analysis. Springer-Verlag New York; 2016.

51. Langfelder P, Horvath S. WGCNA: an R package for weighted correlation network analysis. BMC Bioinformatics. 2008/12/29 2008;9(1):559. doi:10.1186/1471-2105-9-559

52. Knijnenburg TA, Wessels LF, Reinders MJ, Shmulevich I. Fewer permutations, more accurate P-values. Bioinformatics. Jun 15 2009;25(12):i161–8. doi:10.1093/bioinformatics/btp211

53. Szklarczyk D, Gable AL, Nastou KC, et al. The STRING database in 2021: customizable protein-protein networks, and functional characterization of user-uploaded gene/measurement sets. Nucleic Acids Res. Jan 8 2021;49(D1):D605–d612. doi:10.1093/nar/gkaa1074

54. van der Ende EL, In’t Veld S, Hanskamp I, et al. CSF proteomics in autosomal dominant Alzheimer’s disease highlights parallels with sporadic disease. Brain. Nov 2 2023;146(11):4495–4507. doi:10.1093/brain/awad213

55. Baeyens N, Mulligan-Kehoe MJ, Corti F, et al. Syndecan 4 is required for endothelial alignment in flow and atheroprotective signaling. Proceedings of the National Academy of Sciences. 2014;111(48):17308–17313. doi:doi:10.1073/pnas.1413725111

56. Wälchli T, Ghobrial M, Schwab M, et al. Single-cell atlas of the human brain vasculature across development, adulthood and disease. Nature. 2024/08/01 2024;632(8025):603–613. doi:10.1038/s41586-024-07493-y

57. Diaz-Castro B, Bernstein AM, Coppola G, Sofroniew MV, Khakh BS. Molecular and functional properties of cortical astrocytes during peripherally induced neuroinflammation. Cell Rep. Aug 10 2021;36(6):109508. doi:10.1016/j.celrep.2021.109508

58. St-Onge F, Chapleau M, Breitner JCS, Villeneuve S, Pichet Binette A, Initiative ftAsDN. Tau accumulation and its spatial progression across the Alzheimer’s disease spectrum. Brain Communications. 2024;6(1)doi:10.1093/braincomms/fcae031

59. Wojtas AM, Dammer EB, Guo Q, et al. Proteomic Changes in the Human Cerebrovasculature in Alzheimer’s Disease and Related Tauopathies Linked to Peripheral Biomarkers in Plasma and Cerebrospinal Fluid. medRxiv. Jan 11 2024;doi:10.1101/2024.01.10.24301099

60. Zhang GL, Zhang X, Wang XM, Li JP. Towards understanding the roles of heparan sulfate proteoglycans in Alzheimer’s disease. Biomed Res Int. 2014;2014:516028. doi:10.1155/2014/516028

61. Zhu Y, Gandy L, Zhang F, et al. Heparan Sulfate Proteoglycans in Tauopathy. Biomolecules. Nov 30 2022;12(12)doi:10.3390/biom12121792

62. Holmes BB, DeVos SL, Kfoury N, et al. Heparan sulfate proteoglycans mediate internalization and propagation of specific proteopathic seeds. Proceedings of the National Academy of Sciences. 2013;110(33):E3138–E3147. doi:doi:10.1073/pnas.1301440110

63. Rushworth JV, Hooper NM. Lipid Rafts: Linking Alzheimer’s Amyloid-β Production, Aggregation, and Toxicity at Neuronal Membranes. International journal of Alzheimer’s disease. Dec 27 2010;2011:603052. doi:10.4061/2011/603052

64. Hudák A, Kusz E, Domonkos I, et al. Contribution of syndecans to cellular uptake and fibrillation of α-synuclein and tau. Sci Rep. Nov 12 2019;9(1):16543. doi:10.1038/s41598-019-53038-z

65. van Horssen J, Kleinnijenhuis J, Maass CN, et al. Accumulation of heparan sulfate proteoglycans in cerebellar senile plaques. Neurobiol Aging. Jul-Aug 2002;23(4):537–45. doi:10.1016/s0197-4580(02)00010-6

66. Ozsan McMillan I, Li JP, Wang L. Heparan sulfate proteoglycan in Alzheimer’s disease: aberrant expression and functions in molecular pathways related to amyloid-β metabolism. Am J Physiol Cell Physiol. Apr 1 2023;324(4):C893–c909. doi:10.1152/ajpcell.00247.2022

67. Nazere K, Takahashi T, Hara N, et al. Amyloid Beta Is Internalized via Macropinocytosis, an HSPG- and Lipid Raft-Dependent and Rac1-Mediated Process. Front Mol Neurosci. 2022;15:804702. doi:10.3389/fnmol.2022.804702

68. O’Callaghan P, Noborn F, Sehlin D, et al. Apolipoprotein E increases cell association of amyloid-β 40 through heparan sulfate and LRP1 dependent pathways. Amyloid. Jun 2014;21(2):76–87. doi:10.3109/13506129.2013.879643

69. Hussong SA, Banh AQ, Van Skike CE, et al. Soluble pathogenic tau enters brain vascular endothelial cells and drives cellular senescence and brain microvascular dysfunction in a mouse model of tauopathy. Nature Communications. 2023/04/25 2023;14(1):2367. doi:10.1038/s41467-023-37840-y

70. Iadecola C. Untangling Neurons With Endothelial Nitric Oxide. Circ Res. Oct 28 2016;119(10):1052–1054. doi:10.1161/circresaha.116.309927

71. Hoglund Z, Ruiz-Uribe N, del Sastre E, et al. Brain vasculature accumulates tau and is spatially related to tau tangle pathology in Alzheimer’s disease. Acta Neuropathologica. 2024/06/17 2024;147(1):101. doi:10.1007/s00401-024-02751-9

72. Price JL, Ko AI, Wade MJ, Tsou SK, McKeel DW, Morris JC. Neuron number in the entorhinal cortex and CA1 in preclinical Alzheimer disease. Arch Neurol. Sep 2001;58(9):1395–402. doi:10.1001/archneur.58.9.1395

73. Insel PS, Mormino EC, Aisen PS, Thompson WK, Donohue MC. Neuroanatomical spread of amyloid β and tau in Alzheimer’s disease: implications for primary prevention. Brain Commun. 2020;2(1):fcaa007. doi:10.1093/braincomms/fcaa007

74. Tarawneh R, D’Angelo G, Macy E, et al. Visinin-like protein−1: diagnostic and prognostic biomarker in Alzheimer disease. Ann Neurol. Aug 2011;70(2):274–85. doi:10.1002/ana.22448

