## Supplement for "Early Syndecan-4 Upregulation Predicts Cognitive and Pathological Trajectories in Alzheimer Disease"

### Supplementary Tables and Figures

| Supplementary Table 1. CSF Biomarker Characteristics of the Study Cohorts | | | | | | |
|---|---|---|---|---|---|---|
| Study Cohort [†] | All ($n$=1,041) | A+ ($n$=417) | A+T+ ($n$=275) | A- ($n$=624) | Controls ($n$=539) | $P$ value [‡] |
| ***Baseline CSF Biomarker Measures*** [§] | | | | | | |
| SDC4 | 3678 ± 35 | 4053 ± 49 | 4348 ± 58 | 3427 ± 45 | 3324 ± 46 | <0.0001* |
| p-tau181 | 49.6 ± 1.1 | 75.8 ± 2.1 | 94.7 ± 2.4 | 32.2 ± 0.5 | 30.1 ±0.4 | <0.0001* |
| Aβ42/Aβ40 | 0.073 ± 0.001 | 0.047 ± 0.0005 | 0.043 ± 0.0005 | 0.090 ± 0.0004 | 0.090 ± 0.0004 | <0.0001* |
| total tau | 371 ± 8 | 529 ± 14 | 658 ± 16 | 266 ± 7 | 248 ± 7 | <0.0001* |
| VILIP-1 | 2353 ± 34 | 2750 ± 55 | 3161 ± 66 | 2087 ± 39 | 1955 ± 36 | <0.0001* |
| NFL | 5099 ± 116 | 6370 ± 187 | 7299 ± 240 | 4254 ± 138 | 3844 ± 121 | <0.0001* |
| Ng | 4449 ± 42 | 4606 ± 65 | 4786 ± 84 | 4344 ± 55 | 4281 ± 58 | <0.0001* |
| SNAP-25 | 1898 ± 32 | 2116 ± 54 | 2366 ± 67 | 1752 ± 38 | 1671 ± 37 | <0.0001* |
| SYT-1 | 117947 ± 1105 | 126224 ± 1674 | 136979 ± 1946 | 112416 ± 1423 | 108408 ± 1371 | <0.0001* |
| GFAP | 54038 ± 1333 | 68372 ± 2507 | 79940 ± 3413 | 44459 ± 1333 | 40868 ± 1211 | <0.0001* |
| YKL-40 | 803130 ± 10641 | 880856 ± 15866 | 942996 ± 20156 | 751105 ± 13864 | 724831 ± 13076 | <0.0001* |
| sTREM2 | 21248 ± 499 | 24184 ± 914 | 26659 ± 1246 | 19290 ± 552 | 18360 ± 536 | <0.0001* |

*$p$<0.05

† The study cohort ($n$=1,041) included participants who were CDR 0 ($n$=802) and CDR ≥ 0.5 ($n$=239, including CDR 0.5 $n$=186 and CDR 1-2 ($n$=53).

‡ $P$ values reflect comparisons between biomarker-confirmed AD (A+T+, $n$=275) and controls (CDR 0 A-T-; $n$=539)

§ CSF total tau, p-tau181, Aβ42 and Aβ40 (used to measure the Aβ42/Aβ40 ratio) units are in pg/ml. All other biomarker units, including CSF SDC4, are in count values. All values are shown as mean ± standard error.

Abbreviations: CSF, cerebrospinal fluid; SDC4, syndecan-4; p-tau181, tau phosphorylated at threonine 181; Aβ42, amyloid-β peptide 1-42; Aβ40, amyloid-β peptide 1-40; VILIP-1, visinin-like protein-1; NFL, neurofilament-light chain; Ng, neurogranin; SNAP-25, synaptosomal-associated protein-25; SYT-1, synaptotagmin-1; GFAP, glial fibrillary acidic protein; YKL-40, chitinase-3 like protein-1; sTREM2, soluble triggering receptor expressed on myeloid cells-2.

| **Supplementary Table 2. Correlation coefficients of CSF SDC4 with other CSF AD Biomarkers** | | | | |
|---|---|---|---|---|
| | **All (CDR 0 and CDR ≥0.5)** | | **CDR 0** | |
| **CSF Biomarker** | A+ (*n*=417) | A+T+ (*n*=275) | A+ (*n*=228) | A+T+ (*n*=122) |
| p-tau181 † | 0.46 *** | 0.30 *** | 0.48*** | 0.30** |
| Aβ42/Aβ40 † | -0.33 *** | -0.15* | -0.36*** | -0.20* |
| total tau † | 0.46 *** | 0.32*** | 0.48*** | 0.33** |
| VILIP-1 | 0.59 *** | 0.57 *** | 0.63*** | 0.60*** |
| NFL | 0.47 *** | 0.43 *** | 0.41*** | 0.30** |
| Ng | 0.40 *** | 0.35 *** | 0.41*** | 0.34** |
| SNAP-25 | 0.39 *** | 0.36 *** | 0.36*** | 0.28** |
| SYT-1 | 0.50 *** | 0.44 *** | 0.54*** | 0.40*** |
| YKL-40 | 0.58 *** | 0.52 *** | 0.51*** | 0.39*** |
| GFAP | 0.54 *** | 0.52 *** | 0.52*** | 0.43*** |
| sTREM2 | 0.32 *** | 0.28 *** | 0.40*** | 0.43*** |

***$p<0.0001$, ** $p<0.001$, * $p<0.05$

† For CSF p-tau181, Aβ42/Aβ40, and total tau levels, *n*= 412 for the A+ cohort, *n*=272 for the A+T+ cohort, *n*=226 for the CDR 0 A+ cohort, and *n*=121 for the CDR 0 A+T+ cohort.

Abbreviations: CSF, cerebrospinal fluid; SDC4, syndecan-4; p-tau181, tau phosphorylated at threonine 181; Aβ42, amyloid-β peptide 1-42; Aβ40, amyloid-β peptide 1-40; VILIP-1, visinin-like protein-1; NFL, neurofilament-light chain; Ng, neurogranin; SNAP-25, synaptosomal-associated protein-25; SYT-1, synaptotagmin-1; GFAP, glial fibrillary acidic protein; YKL-40, chitinase-3 like protein-1; sTREM2, soluble triggering receptor expressed on myeloid cells-2.

| **Supplementary Table 3. Correlations of CSF SDC4 levels with Global and Braak stages of Tau-PET Burden**[†] | | | | |
|---|---|---|---|---|
| | Combined ($n$=302) | Aβ+ ($n$=92) | CDR 0-0.5 ($n$=293) | CDR 0-0.5 Aβ+ ($n$=84) |
| Global Tau | 0.25 ($p<0.0001$)* | 0.34 ($p=0.001$)* | 0.25 ($p<0.0001$)* | 0.35 ($p=0.001$)* |
| Braak 1-2 | 0.20 ($p<0.0001$)* | 0.21 ($p=0.04$)* | 0.17 ($p=0.004$)* | 0.19 ($p=0.06$) |
| Braak 3-4 | 0.19 ($p<0.0001$)* | 0.25 ($p=0.017$)* | 0.18 ($p=0.002$)* | 0.27 ($p=0.014$)* |
| Braak 5-6 | 0.22 ($p<0.0001$)* | 0.30 ($p=0.004$)* | 0.25 ($p<0.0001$)* | 0.36 ($p<0.0001$)* |

*$p<0.05$

[†] Values represent partial correlation coefficients adjusting for age and sex. *P* values are shown in parentheses.

Abbreviations: CSF, cerebrospinal fluid; SDC4, syndecan-4; CDR, Clinical Dementia Rating.

| **Supplementary Table 4. CSF SDC4 Correlations with Regional Estimates of Tau-PET Burden[†]** | | |
|---|---|---|
| **Brain region** | **All (*n*=302)** | **Tau-PET positive (*n*=135)** |
| Banks of Superior Temporal Sulcus | **0.19 (*p*<0.0001) *** | **0.24 (*p*=0.005) *** |
| Caudal Middle Frontal | 0.18 (*p*=0.002) * | 0.22 (*p*=0.009) * |
| Entorhinal | **0.20 (*p*<0.0001) *** | **0.23 (*p*=0.009) *** |
| Parahippocampal | 0.15 (*p*=0.009) * | 0.18 (*p*=0.03) * |
| Fusiform | **0.21 (*p*<0.0001) *** | **0.28 (*p*=0.001) *** |
| Inferior Temporal | **0.24 (*p*<0.0001) *** | **0.33 (*p*<0.0001) *** |
| Middle Temporal | **0.25 (*p*<0.0001) *** | **0.33 (*p*<0.0001) *** |
| Superior Temporal | **0.20 (*p*<0.0001) *** | **0.23 (*p*=0.008) *** |
| Temporal Pole | **0.20 (*p*<0.0001) *** | **0.33 (*p*<0.0001) *** |
| Posterior Cingulate | 0.15 (*p*=0.008) * | 0.17 (*p*=0.05) * |
| Isthmus of Cingulate | **0.19 (*p*<0.0001) *** | **0.25 (*p*=0.003) *** |
| Precuneus | 0.17 (*p*=0.004) * | 0.22 (*p*=0.01) * |
| Inferior Parietal | **0.24 (*p*<0.0001) *** | **0.31 (*p*<0.0001) *** |
| Supramarginal | **0.21 (*p*<0.0001) *** | **0.24 (*p*=0.005) *** |
| Lateral Occipital | 0.16 (*p*=0.005) * | 0.21 (*p*=0.02) * |
| Superior Parietal | 0.13 (*p*=0.03) * | 0.16 (*p*=0.06) |
| Superior Frontal | 0.13 (*p*=0.03) * | 0.15 (*p*=0.07) |
| Insula | 0.18 (*p*=0.001) * | 0.20 (*p*=0.02) * |
| Lateral Orbitofrontal | 0.12 (*p*=0.045) * | 0.12 (*p*=0.17) |
| Medial Orbitofrontal | 0.13 (*p*=0.02) * | 0.09 (*p*=0.28) |
| Rostral Middle Frontal | 0.14 (*p*=0.01) * | 0.17 (*p*=0.05) * |
| Lingual | 0.11 (*p*=0.06) | 0.12 (*p*=0.16) |
| Pars Opercularis | 0.11 (*p*=0.06) | 0.17 (*p*=0.06) |

*$p$<0.05
[†] Values represent partial correlation coefficients adjusting for age and sex. *P* values are shown in parentheses.
No significant correlations were observed between CSF SDC4 and regional tau burden in the caudal anterior cingulate, rostral anterior cingulate, cuneus, frontal pole, precentral, paracentral, postcentral, pars orbitalis, pars triangularis, pericalcarine, or transtemporal regions in the combined or tau-PET positive cohorts (data not shown). Regions known to be involved in early AD are shown in bold.

| **Supplementary Table 5. CSF SDC4 Correlations with Regional Amyloid Burden (PiB only; *n*=528)** | |
|---|---|
| **Brain region** | **Correlation coefficient** † |
| Banks of Superior Temporal Sulcus | 0.16 ($p$<0.0001) * |
| Caudal Anterior Cingulate | 0.15 ($p$<0.0001) * |
| Caudal Middle Frontal | 0.13 ($p$=0.003) * |
| Entorhinal | 0.13 ($p$=0.004) * |
| Parahippocampal | 0.12 ($p$=0.005) * |
| Fusiform | 0.17 ($p$<0.0001) * |
| Inferior Temporal | 0.17 ($p$<0.0001) * |
| Middle Temporal | 0.16 ($p$<0.0001) * |
| Superior Temporal | 0.17 ($p$<0.0001) * |
| Temporal Pole | 0.13 ($p$=0.003) * |
| Transtemporal | 0.17 ($p$<0.0001) * |
| Rostral Anterior Cingulate | 0.17 ($p$<0.0001) * |
| Posterior Cingulate | 0.14 ($p$=0.001) * |
| Isthmus of Cingulate | 0.12 ($p$=0.004) * |
| Precuneus | 0.14 ($p$=0.001) * |
| Inferior Parietal | 0.14 ($p$<0.0001) * |
| Supramarginal | 0.17 ($p$<0.0001) * |
| Lateral Occipital | 0.10 ($p$=0.01)* |
| Superior Parietal | 0.12 ($p$=0.007) * |
| Frontal Pole | 0.15 ($p$<0.0001) * |
| Superior Frontal | 0.15 ($p$<0.0001) * |
| Insula | 0.16 ($p$<0.0001) * |
| Lateral Orbitofrontal | 0.16 ($p$<0.0001) * |
| Medial Orbitofrontal | 0.15 ($p$<0.0001) * |
| Rostral Middle Frontal | 0.16 ($p$<0.0001) * |
| Lingual | 0.12 ($p$=0.004) * |
| Pars Orbitalis | 0.16 ($p$<0.0001) * |
| Pars Opercularis | 0.18 ($p$<0.0001) * |
| Pars Triangularis | 0.18 ($p$<0.0001) * |
| Precentral | 0.14 ($p$=0.001) * |
| Paracentral | 0.14 ($p$=0.001) * |
| Postcentral | 0.14 ($p$=0.002) * |

*$p$<0.05

† Values represent partial correlation coefficients adjusting for age, sex, and *APOE ε4*. *P* values are shown in parentheses.

| **Supplementary Table 6. CSF SDC4 Correlations with Baseline Cognitive Outcomes** † | | | | | | |
|---|---|---|---|---|---|---|
| **Cognitive Measure** | Combined (all CDR) (*n*=1,041) | A+ (all CDR) (*n*=417) | CDR 0-0.5 (*n*=988) | A+ (CDR 0-0.5) (*n*=367) | CDR 0 (*n*=802) | A+ CDR 0 (*n*=228) |
| **CDR-SB** | 0.15 ($p$<0.0001) * | 0.13 ($p$=0.007) * | 0.17 ($p$<0.0001) * | 0.15 ($p$=0.004) * | - | - |
| **Knight-PACC**‡ | -0.18 ($p$<0.0001) * | -0.20 ($p$<0.0001) * | -0.18 ($p$<0.0001) * | -0.23 ($p$<0.0001) * | -0.14 ($p$<0.0001) * | -0.21 ($p$=0.002) * |
| **FCSRT-Free**‡ | -0.15 ($p$<0.0001) * | -0.13 ($p$=0.01) * | -0.15 ($p$<0.0001) * | -0.16 ($p$=0.004) * | -0.11 ($p$=0.002) * | -0.16 ($p$=0.02) * |
| **Animal Fluency**‡ | -0.13 ($p$<0.0001) * | -0.20 ($p$<0.0001) * | -0.12 ($p$<0.0001) * | -0.20 ($p$<0.0001) * | -0.09 ($p$=0.01) * | -0.20 ($p$=0.003) * |
| **Digit-Symbol**‡ | -0.04 ($p$=0.28) | -0.05 ($p$=0.35) | -0.02 ($p$=0.58) | -0.05 ($p$=0.44) | 0.042 ($p$=0.35) | -0.01 ($p$=0.90) |
| **Trail-Making B**‡ | 0.01 ($p$=0.77) | 0.04 ($p$=0.45) | -0.02 ($p$=0.61) | 0.02 ($p$=0.72) | -0.07 ($p$=0.06) | 0.01 ($p$=0.91) |

*$p$<0.05

† Values represent partial correlation coefficients of CSF SDC4 with cognitive outcomes, adjusting for age, sex, education, and *APOE ε4.* CSF SDC4 levels and all cognitive outcomes were standardized to $z$ scores prior to analysis.

‡ $n$=1,018 for the combined (CDR 0 and CDR ≥ 0.5), $n$=408 for the A+, $n$=970 for the CDR 0-0.5, $n$=363 for the A+ CDR 0-0.5, $n$=790 for the CDR 0, and $n$=227 for the A+ CDR 0 cohorts.

*P* values are shown in parentheses.

**Supplementary Figure 1**

**A** **B**

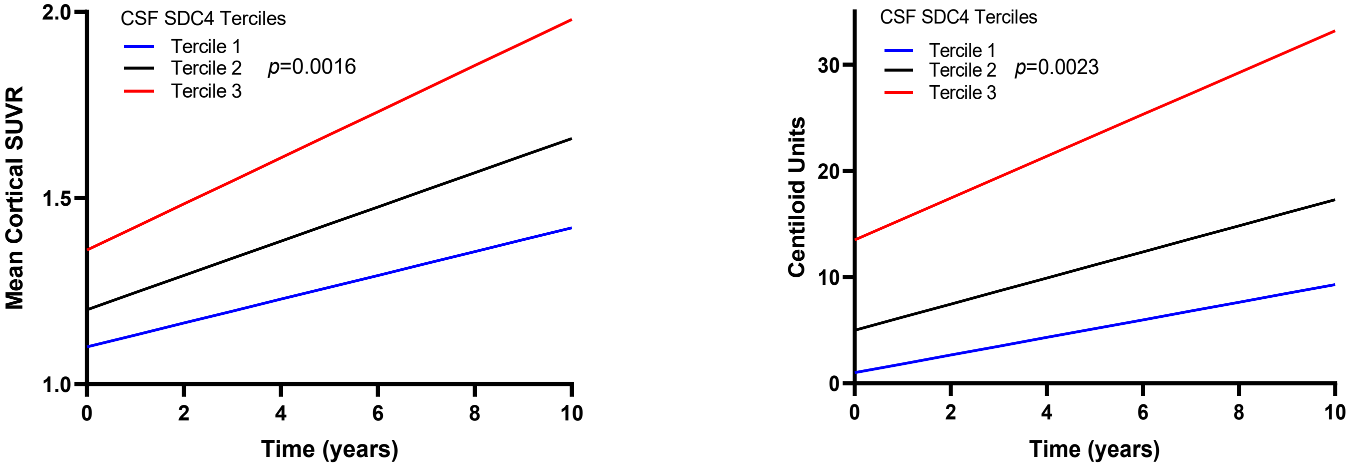


**Supplementary Figure 1. Adjusted annual rates of amyloid-PET progression as a function of CSF SDC4 levels.** Participants ($n$=370) with longitudinal amyloid-PET scans whose baseline CSF SDC4 levels were in the upper tercile demonstrated more rapid progression in global amyloid burden, measured using **(A)** the (untransformed) mean cortical SUVR ($p$=0.0016) or **(B)** Centiloid Units ($p$=0.0023), over time compared to those whose SDC4 levels were in the middle or lower terciles, adjusting for age, sex, and *APOE ε4*. Adjusted rates of progression in mean (untransformed) cortical SUVR values were 0.032 ± 0.005, 0.046 ± 0.006, and 0.062 ± 0.006, respectively for the lower, middle, and upper terciles. Adjusted rates of progression in Centiloid Units were 0.83 ± 0.22, 1.23 ± 0.23, and 1.97 ± 0.25, respectively for the lower, middle, and upper terciles. Cut-off values for CSF SDC4 levels were: 2974 ($33^{rd}$ percentile) and 3678 ($67^{th}$ percentile). The shown intercepts are model estimates adjusting for age, sex, and *APOE ε4*.

**Supplementary Figure 2**

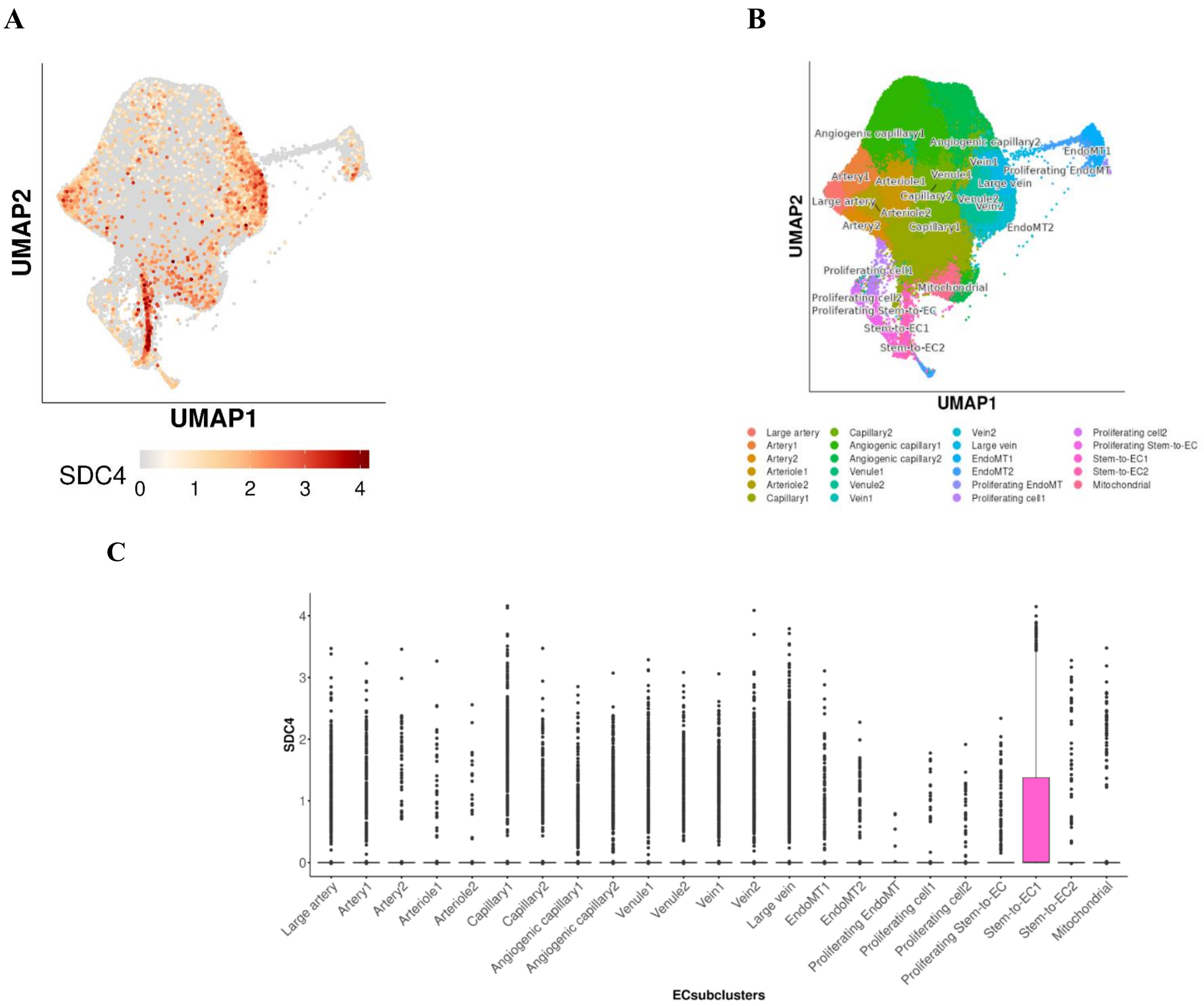


**Supplementary Figure 2. Brain Endothelial expression of SDC4. (A)** SDC4 expression in clusters representing endothelial cell subtypes obtained from a single-cell atlas of the human brain vasculature which examined CD31+/CD45- FACS (fluorescence-activated cell sorting)-sorted brain endothelial cells. SDC4 is mostly expressed in the early stages of endothelial cell development (i.e., stem cells to endothelial cells) and proliferating endothelial cells. **(B)** Color scheme representing the brain endothelial cell subclusters (i.e., subtypes) obtained from a human brain vascular atlas. **(C)** A box-plot representing the expression of SDC4 ( i.e. the number of unique molecular identifiers [UMIs] per cell) in brain endothelial subtypes. The highest SDC4 expression levels were observed in endothelial cells developing from stem cells (i.e., during angiogenesis). These figures were generated using an interactive single-cell atlas of the human brain vasculature available at Wälchli T. et al.[1]

**Supplementary Figure 3**

**A** **B**

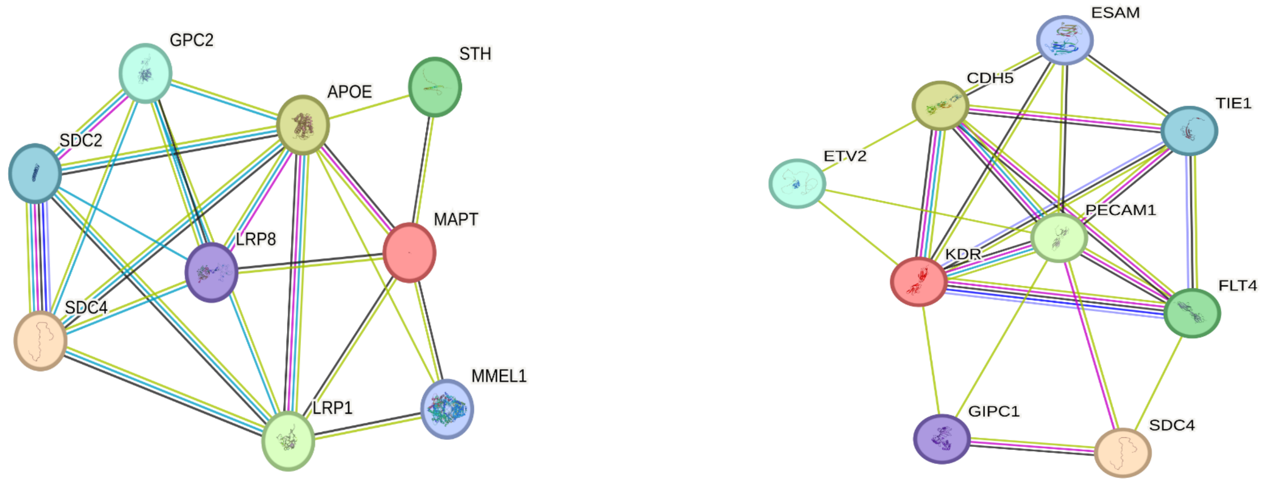


**Supplementary Figure 3.** Functional pathway analyses using STRING. **(A)** Functional pathway analyses demonstrate interactions of *SDC4* with AD-related proteins including *APOE*, *LRP1*, *LRP8, and MAPT*. The combined evidence score (suggesting a functional link) for the *SDC4–APOE* interaction is 0.77, for *SDC4-LRP1* is 0.48, and for *SDC4-LRP8* is 0.41. **(B)** Functional pathway analyses demonstrate interactions of *SDC4* with other endothelial proteins, including *PECAM1*, *FLT4*, and *GIPC1*. The combined evidence score (suggesting a functional link) for *SDC4-PECAM1* is 0.63, for *SDC4-GIPC1* is 0.97, and *SDC4-FLT4* is 0.50. Network nodes represent proteins; splice isoforms or post-translational modifications are collapsed so that each node represents all the proteins produced by a single protein-coding gene locus. Edges represent protein–protein associations that are meant to be specific and meaningful (i.e., proteins jointly contribute to a shared function) and do not necessarily mean the proteins are physically binding. Color coding for the interactions are as follows: known interactions from curated databases (teal); known experimentally determined interactions (purple); predicted interactions from gene neighborhood (green), gene fusions (red), or gene co-occurrence (blue); predicted interactions from text-mining (yellow), co-expression (black), and protein homology (lavender). Abbreviations, *APOE*, apolipoprotein E; *CDH5*, cadherin-5 (aka vascular endothelial-cadherin); *ESAM*, endothelial cell-selective adhesion molecule; *ETV2*, ETS translocation variant 2; *FLT4*, vascular endothelial growth factor receptor 3; *GIPC1*, GAIP-interacting protein, C terminus 1 (aka PDZ domain-containing protein GIPC1); *GPC2*, secreted glypican-2; *KDR*, vascular endothelial growth factor receptor 2; *LRP*, low-density lipoprotein receptor-related protein; *MAPT*, microtubule-associated protein; *MMEL1*, membrane metallo-endopeptidase-like 1; *PECAM*, platelet-endothelial cell adhesion molecule; *SDC4*, syndecan-4; *STH*, saitohin; *TIE1*, tyrosine-protein kinase receptor Tie-1.

**References:**

1. Wälchli T, Ghobrial M, Schwab M, et al. Single-cell atlas of the human brain vasculature across development, adulthood and disease. *Nature*. 2024/08/01 2024;632(8025):603-613. doi:10.1038/s41586-024-07493-y
